# Maturity Onset Diabetes of the Young (MODY) is not always monogenic: candidate genes involved in a Latino Population

**DOI:** 10.1101/2024.10.02.24314794

**Authors:** Alberto Moscona-Nissan, Daniel Marrero-Rodríguez, Sergio Andonegui-Elguera, Eduardo Salif Luna-Ávila, Florencia Martínez-Mendoza, Sandra Vela-Patiño, Itzel Ramírez-Ramos, Silvia Hinojosa-Alvarez, Jesus Hernandez-Perez, Rocio A Chavez-Santoscoy, Sophia Mercado-Medrez, Kapy S León-Wu, Regina De Miguel-Ibáñez, Moisés Mercado, Keiko Taniguchi-Ponciano, Aldo Ferreira-Hermosillo

## Abstract

**Introduction:** MODY misdiagnosis remains widespread, existing remarkable variability within genetic variants across populations. While diagnostic tools are based on Caucasian cohorts, Whole Exome Sequencing (WES) studies are needed to identify new genes in non-Caucasians, as up to 77% of patients do not harbor variants of significance in MODY-known genes. In Latino populations, no WES studies have addressed MODY genomic landscape beyond its canonical genes.

**Methods:** We carried out WES in 17 patients with MODY, 17 patients with type 2 diabetes (T2DM) and 17 healthy controls (HC). MODY diagnosis was established according to Exeter criteria (score ≥36%) in subjects with no or minimal insulin requirements. We compared the single nucleotide variant (SNV) landscape across groups.

**Results:** Patients with MODY present a polygenic landscape with allelic variants in canonical and non-canonical genes. Canonical MODY genes used for routine genetic diagnosis showed low discrimination utility, having similar frequencies between MODY, T2DM and HC in the Mexican population. We propose 14 genes with variants that distinguish MODY from T2DM and HC, as we detected variants in genes as *MAP2K3, SYT15, TPTE, KCNJ12, PEX5,* and *OR2A1* in 75-100% of MODY cases while were absent in T2DM and HC. Enrichment analysis revealed involvement in synaptic vesicle trafficking, insulin/IGF pathway-mitogen activated protein kinase kinase/MAPK cascade, and insulin/IGF pathway-protein kinase B/AKT signaling cascade.

**Discussion:** MODY presents a polygenic landscape. Besides improving our understanding of glycemic regulation pathways, the candidate genes could serve as MODY diagnostic biomarkers in Latino populations.

**Funding:** Supported by grant R-2019-785-052 from Instituto Mexicano del Seguro Social.

## Introduction

Maturity-onset diabetes of the young (MODY) comprises a clinically heterogeneous group of monogenic forms of diabetes characterized by autosomal dominant mode of inheritance, early onset, and absence of pancreatic beta-cell autoimmunity.^1^ Up to 14 types of MODY have been described, related to genes with key roles in pancreatic beta-cell differentiation, insulin secretion, and glucose metabolism.^2^ The most frequently affected genes include those encoding hepatocyte nuclear factor 1α (*HNF1A*), hepatocyte nuclear factor 4α (*HNF4A),* and glucokinase (*GCK*). Meanwhile, mutations in other genes (*PDX1, HNF1B*, *NEUROD1*, *KLF11*, *CEL*, *PAX4*, *INS*, *BLK*, *ABCC8, KCNJ11,* and *APPL1*) have been described but are rather infrequent.^3^

It is estimated that only 3% of MODY patients are diagnosed correctly, while the rest are mistakenly classified as type 1 (T1DM) or type 2 diabetes mellitus (T2DM) in 36% and 51% of cases, respectively.^4–5^ Misdiagnosis of MODY remains widespread, as physicians face challenges such as overlapping clinical phenotypes between MODY and other types of diabetes.^6^ Establishing an accurate diagnosis is of paramount importance to offer tailored therapies, genetic counseling, and prevent complications.^6^

The remarkable variability within genetic variants in MODY patients across world populations, along with reliance on clinical and genetic diagnostic tools based on European and Caucasian cohorts, prompt the need of whole exome sequencing (WES) studies to identify new genes involved in MODY in non-Caucasian populations.^7^ This knowledge gap is highlighted by the fact that up to 77% of Middle Eastern patients with MODY lack variants of significance in known-MODY genes,^8^ while other populations have genetic confirmation rates as low as 6.6%.^9^ In the Latino population, no WES studies have addressed MODY genetic landscape beyond its canonical genes.

Furthermore, studying the genetic aspects of diabetes is crucial in Latino populations as they present a disproportionately heavy burden imposed by this condition, because of its increased prevalence, alongside underrepresentation in DNA-based studies.^10–11^ In Mexico, despite the high prevalence of diabetes (18.3%) associated with the even higher prevalence of overweight (38.3%) and obesity (36.9%), few studies have addressed MODY-related genetic factors.^12^

The aim of this study was to analyze and characterize the genetic variants present in Mexican patients with MODY through WES, comparing them to a cohort of patients with recent diagnosis of T2DM, and young healthy controls (HC).

## Methods

### Clinical features, selection criteria and definitions

We enrolled 51 participants divided in three groups (MODY, T2DM and HC), each having 17 subjects. All MODY and T2DM patients were treated and followed at Centro Médico Nacional Siglo XXI of the Instituto Mexicano del Seguro Social, the largest tertiary care facility in Mexico. All subjects signed an informed consent. The study protocol was approved by the scientific and ethics committees (R-2023-3601-003 and R-2023-3601-212) and conducted in accordance with clinical research laws and the Helsinki declaration.

Inclusion criteria for MODY included adult subjects (≥18 years), with an Exeter score ≥36%, low (<1 U/Kg/ day) or null insulin requirements, preserved beta-cell function (serum C-peptide >1ng/mL) and lack of T1DM autoantibodies. The T2DM group included adult patients with <10 years of diagnosis prior to enrollment, with any type of treatment, regardless of HbA1c levels. HC comprised adult subjects, without parental history of diabetes or metabolic diseases such as hypertension or dyslipidemia. We retrieved clinical, biochemical, and anthropometric information from medical records and calculated the Exeter score for MODY and T2DM cases diagnosed under 35 years, available at: https://www.diabetesgenes.org/exeter-diabetes-app/.

### Whole exome sequencing (WES)

Genomic DNA (gDNA) was extracted from peripheral blood mononuclear cells using the Qiagen kit protocol. DNA purification was carried out with the DNAeasy (Qiagen Inc, CA, USA) blood and tissue kit. Leukocytes were lysed using proteinase-K and the cell lysate was subsequently transferred to the DNAeasy columns. Once DNA was captured in columns, washes were carried out with the kit’s specific buffers to perform DNA elution with molecular biology grade water. High quality, high molecular weight DNA was determined using the NanoDrop2000 and the TapeStation (Agilent Technologies, CA, USA).

The gDNA was shipped to the Genomics Core Lab of the Tecnológico de Monterrey for WES. gDNA was quantified using Qubit dsDNA BR Assay Kit (Invitrogen, Carlsbad, CA, USA). Quality was determined spectrophotometrically using Nanodrop One (Thermo Fisher Scientific, Waltham MA, USA). WES libraries were prepared using Illumina DNA Prep with Exome 1.0 Enrichment (Illumina, San Diego CA, United States) and quantified with Qubit dsDNA BR Assay Kit (Invitrogen, Carlsbad, CA, USA), library size was analyzed in S2 Standard DNA Cartridge for Sep 400 (BiOptic, Taiwan), and sequencing was performed in NovaSeq6000 (Illumina, San Diego CA, USA) in a 150 bp paired-end configuration.

### Computational bioinformatic and statistical analysis

Preprocessed sequences were aligned to human reference sequence (hg38) using the Illumina-Dragen Enrichment pipeline (llumina, San Diego CA, USA). This pipeline was set to produce copy number variants (enable-cnv true). The BAM files resulting from the enrichment were removed from PCR duplicates using Picard Tools (http://broadinstitute.github.io/picard). Each BAM file was used to obtain the somatic variants using the GATK pipeline, and variants where annotated using ANNOVAR according to the following databases: Clinvar, ClinSig, SIFT, Polyphen-2 (HUMVAR-2), gnomAD, refGene, cytoBand, exac03, avsnp147, and dbnsfp30a.

VCF files were transformed and annotated using the vcf2maf tool and Variant Effect Predictor (VEP) 112 release database of ENSEMBL. In addition, the Maftools package version 2.21.1 in R language version 4.4.1 was used to analyze and visualize the landscape of variants. Groups were constructed using read.maf and merge_maf functions. To compare groups mafCompare function was used with a Fisher test on a 2×2 contingency table and to detect differences across genes in groups. Graphs of mutations in genes of interest were constructed using Lollipop Plots to observe mutations by study subgroup using cBioPortal (www.cbioportal.org/). We considered genetic variants with a frequency >75% across MODY patients and absent in T2DM and HC and used Web-gestalt for enrichment analysis (www.webgestalt.org).

We filtered all MODY 1-14 genetic variants to evaluate variant frequency in every patient and group. We tracked an additional set of genes reported in the literature for their involvement in MODY (*RFX6*, *WFS1*, *NKX6-1*, *AKT2*, *NKX2-2*, *PCBD1*, *MTOR*, *TBC1D4*, *CACNA1E*, and *MNX1*) and the specific variants of genes with significant differences across groups through Fisher test. We carried genetic analysis upon individualized cases to explore the association of variants per MODY case. We constructed a clustered heatmap on ClustVis (version 0.7.7) of MODY 1-14 variants. Comparisons of genetic variant frequency across groups were conducted using Chi-square test. T-Student test, and one-way ANOVA were also used when applicable for analyzing sociodemographic data. A p-value <0.05 was considered significant. Statistical package consisted of SPSS 24.0 (IBM, New York, USA).

## Results

### Baseline characteristics

A total of 51 patients were included in our study across 3 groups (MODY, T2DM, and HC) each comprising 17 subjects. The mean age at evaluation of MODY patients was 21.2 ± 5.7 years, 43.5 ± 10.1 years in T2DM patients, and 25.5 ± 3.4 years in HC. Mean age at diabetes diagnosis was significantly lower in the MODY group, being 21.2 years, compared to 43.5 years in T2DM group (p<0.001). No significant differences among groups were found regarding sex. Parental history of diabetes was present in 88% of MODY patients, and in 53% of T2DM subjects (p=0.023).

Considering the clinical presentation of MODY cases, type 3 MODY (*HNF1a*) was the most frequently suspected subtype in 47% (8/17) of patients, followed by dual suspicion of MODY 3 (*HNF1a*)/MODY 5 (*HNF1B*) in 22% of cases, MODY 9 (*PAX4* in ketosis prone patients) suspicion in 11% of patients, and MODY 2 (*GCK*), MODY 5 (*HNF1B*), and MODY2/5 dual suspicion in single cases (Table 1).

**Table 1.**
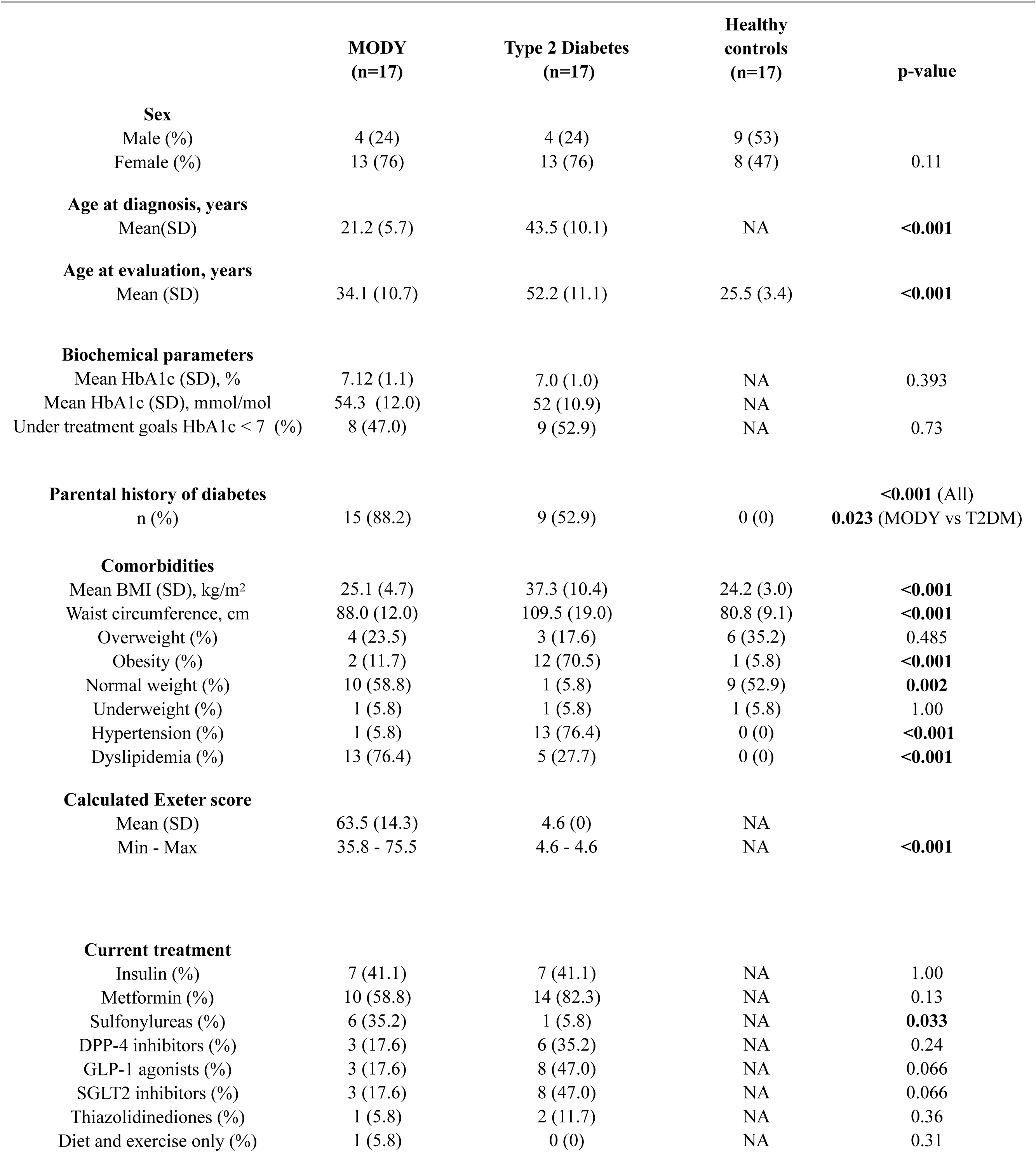
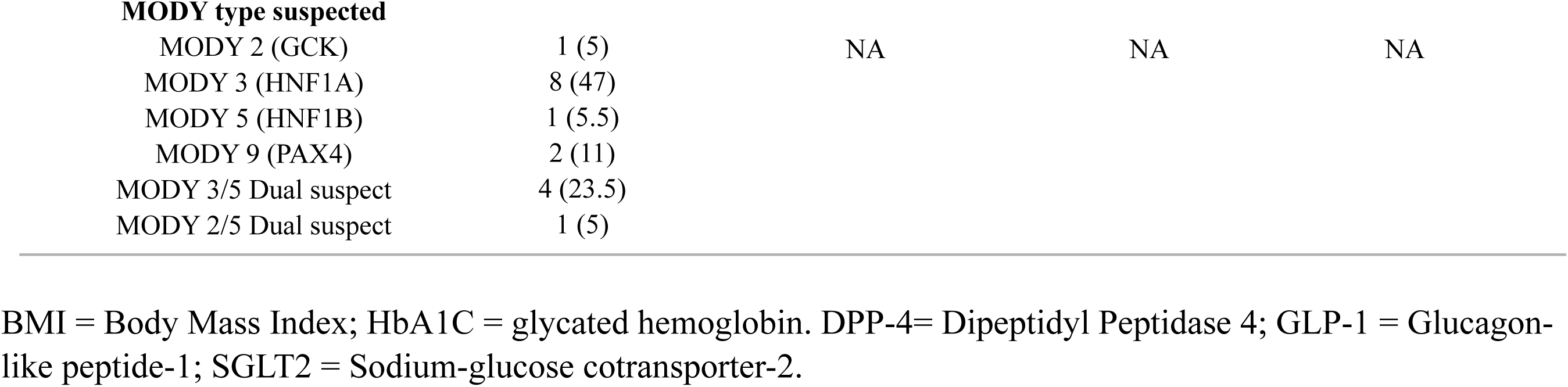
Sociodemographic and clinical data of included participants.

Mean HbA1c levels were similar in MODY and T2DM patients (7.1% and 7.0%, respectively), as 47% and 53% of patients reached treatment goals, respectively. BMI was significantly higher among patients with T2DM. Upon evaluation of comorbidities, the T2DM group exhibited the highest frequency of hypertension, whereas dyslipidemia was more prevalent among MODY patients. The mean Exeter score in MODY patients was 63.5 ± 14.3 (35.8-75.5%), while patients with T2DM had a score of 4.6%. The only difference between groups regarding treatment was that >35% of MODY patients were treated with sulfonylureas, versus 5.5% of T2DM cases.

### WES analysis of MODY, T2DM and HC reveals low utility of canonical diagnostic genes

All groups exhibited similarities in the overall variant distribution. However, each group presented genes with unique variants. MODY, T2DM, and HC displayed an average range of 11,057 to 11,822 variants, with missense variants being the most prevalent. We found 92 distinct genetic variants among the 14-known MODY genes across all subjects. *ABCC8, CEL, BLK,* and *HNF1A* presented the highest burden of variants, while no variants were detected in the *INS* gene (MODY 10), as displayed in Supplement Table 1 and Figure 1.

**Figure 1.**
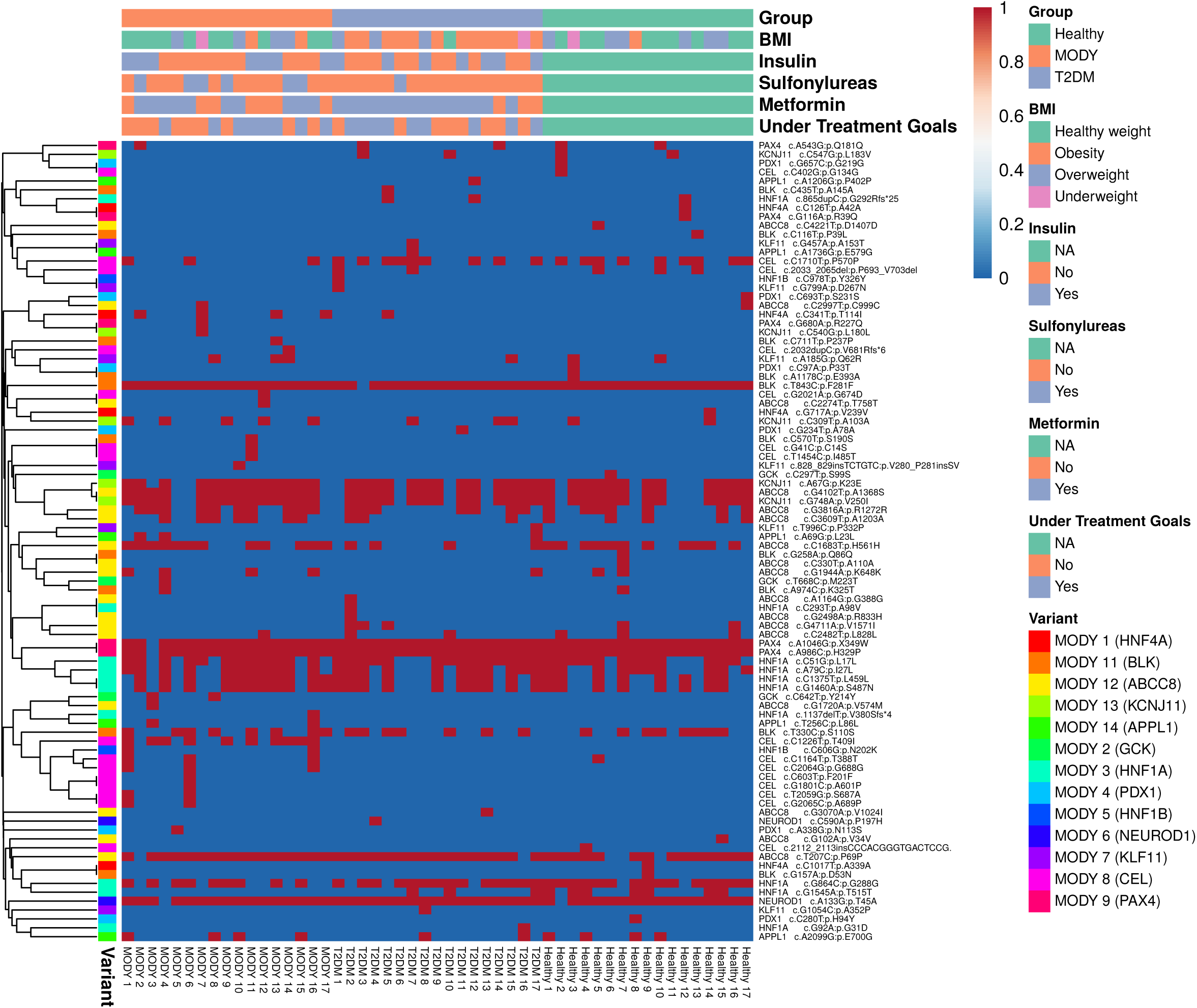
Clustered heatmap of genetic variants for MODY 1-14 genes detected across groups. BMI=Body Mass Index; Under Treatment Goals criteria was defined as a glycated hemoglobin equal or lower than 7.0%.

**Figure 2.**
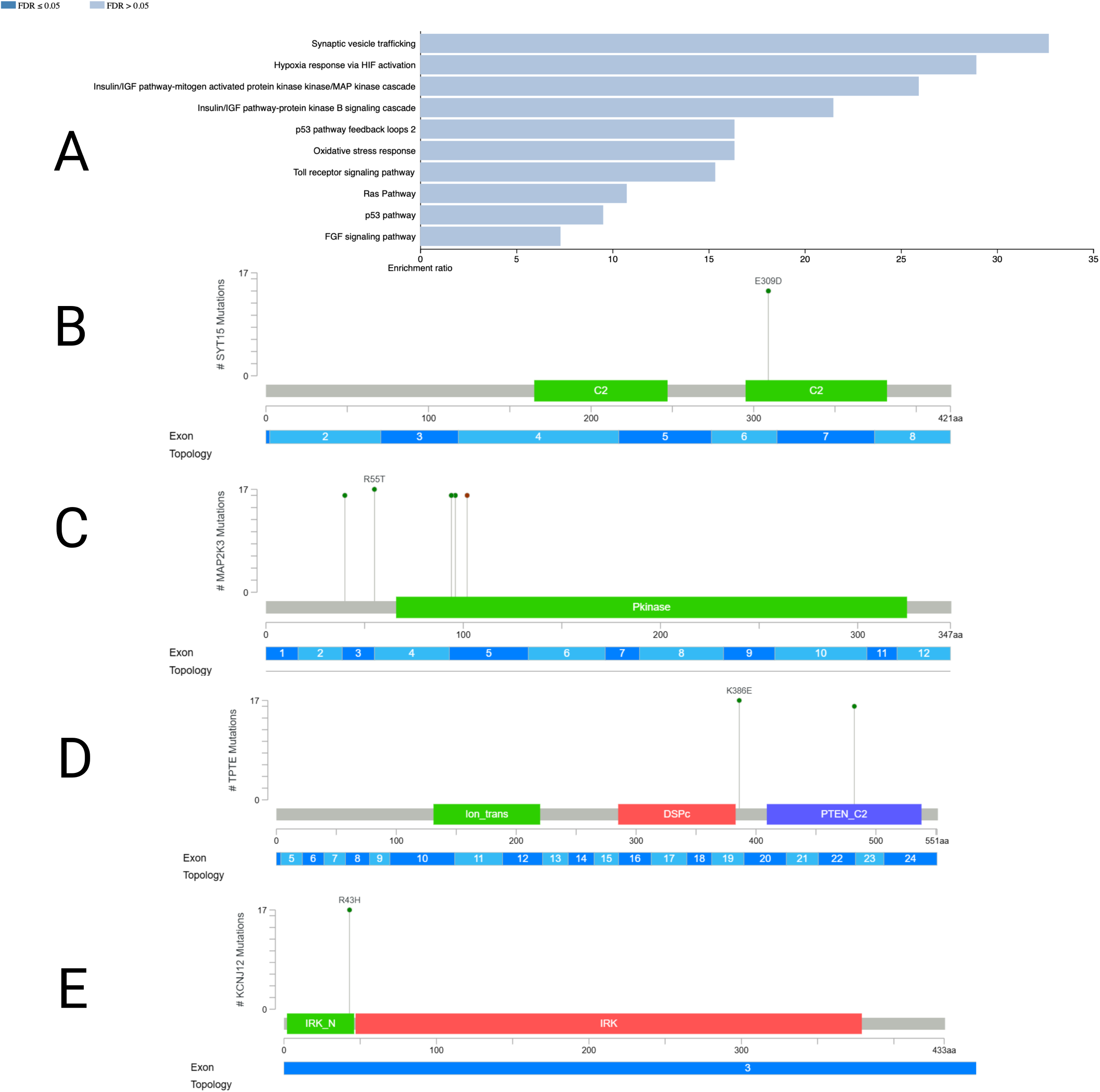
Enrichment analysis of MODY candidate genes highlights key pathways of glucose metabolism (A) and lollipop diagrams of MODY-enriched variants in SYT15 (B), MAP2K3 (C), TPTE (D), and KCNJ12 (E) genes.

Interestingly, most MODY patients showed several variants in the same gene used for diagnosis (i.e. HNF1A) in addition to allelic variants in other genes used for the diagnosis of a different MODY subtype (i.e. *NEUROD1*), rendering the genetic diagnosis rather difficult. Many cases of MODY type 3 suspects showed several allelic variations in *HNF1A* alongside with genetic variants in *CEL, NEUROD1* and *PAX4* (Figure 1). Moreover, most variants in genes canonically used for MODY diagnosis are present in both, T2DM and HC (Figure 1, supplementary table 1). Therefore, the use of canonical or previously described genes used for MODY diagnosis, in Mexican population does not seem appropriate.

When comparing MODY group with T2DM and HC, we only found statistically significant differences in genes such as *CEL* particularly in the variant c.C1226T:p.T409I (p=0.001), which was present in 10 of 17 MODY cases, although *CEL* gene is used for MODY type 8 diagnosis, and was present in patients with MODY type 3 suspect, while it was absent in T2DM and HC groups.

We analyzed other genes associated with MODY in other populations (*RFX6, WFS1, NKX6-1, AKT2, NKX2-2, PCBD1, MTOR, TBC1D4, CACNA1E, and MNX1*) and did not find any significant differences regarding variant frequency among groups (Supplement Table 2).

### WES analysis of MODY, T2DM and HC reveals potential differential diagnostic genes

We searched for differential genetic variant enrichment across groups. We identified 14 genes with variants detected in 75-100% of MODY patients, that were absent in the rest of groups,. These genes include *MAP2K, TPTE, SYT15, PEX5, KCNJ12, KTM2C, OR2A1, RIMBP3, TRIM49C, AQP12B, OR51A4, RIMBP3B, ZNF717,* and *SUSD2*. In total, we detected 27 variants across these genes, which we describe in detail in Table 3.

**Table 2.**
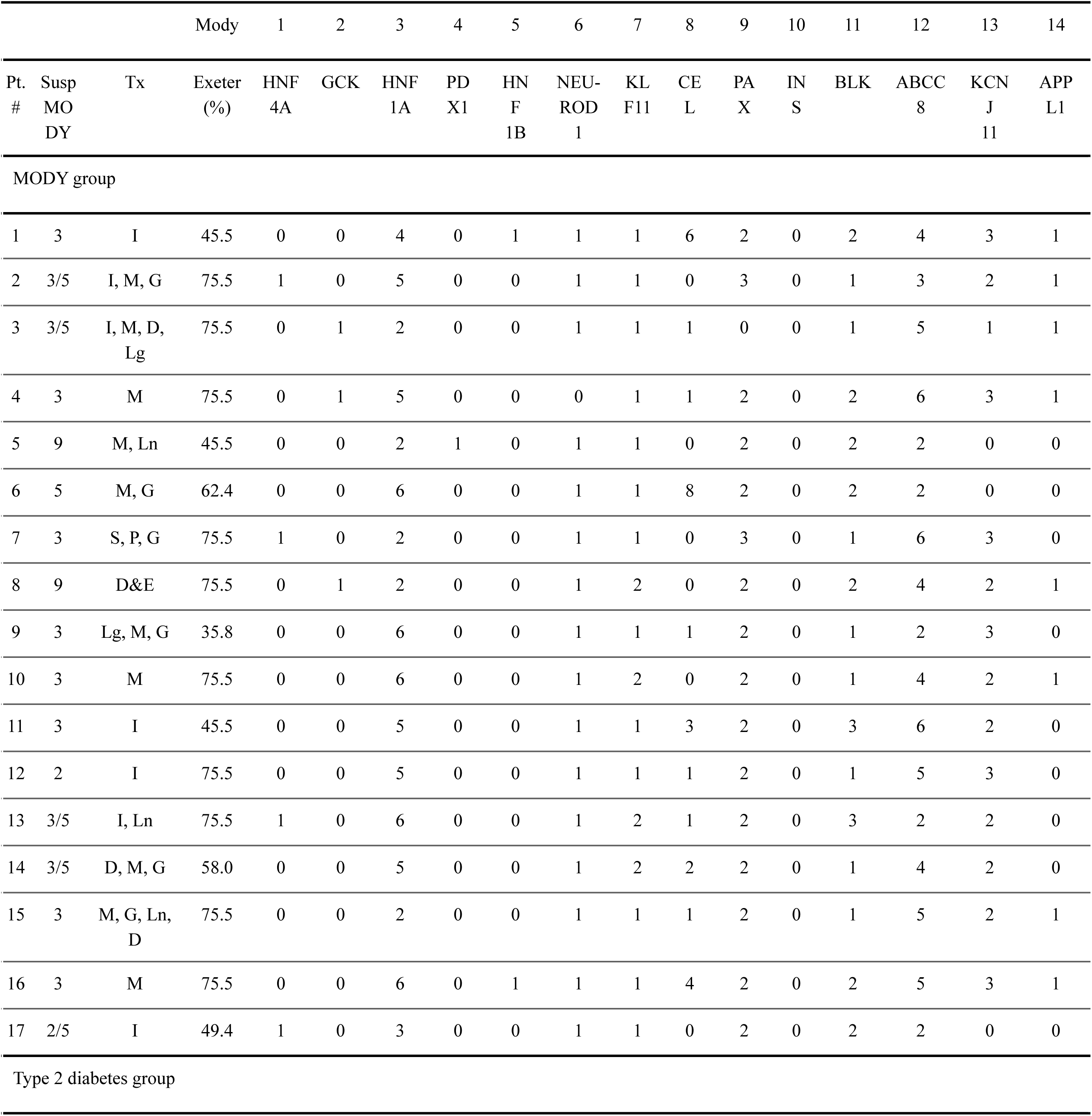
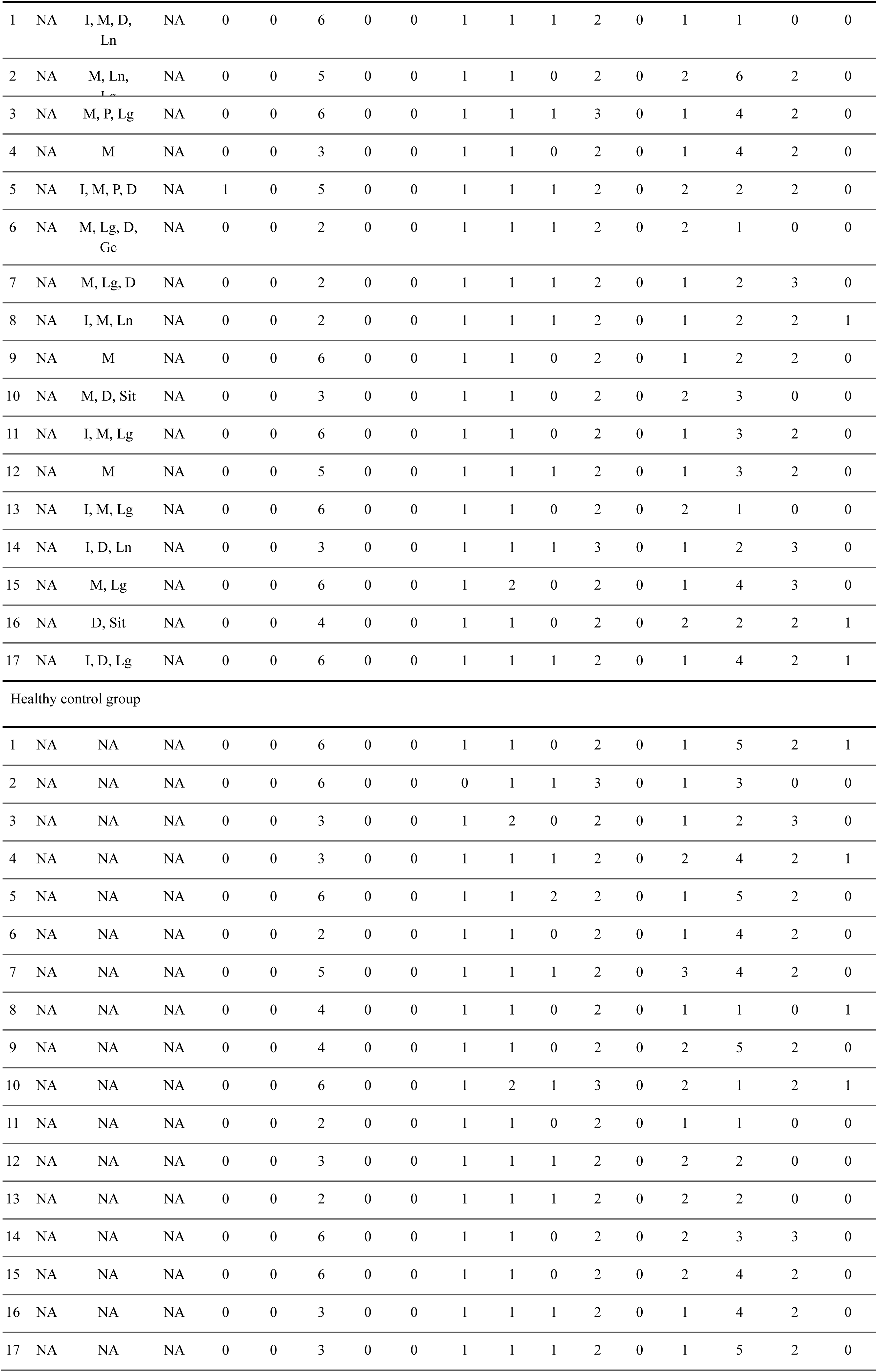
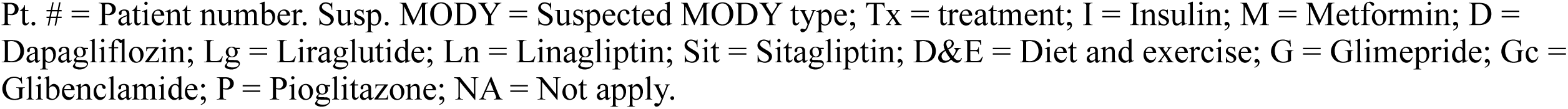
Number of detected genetic variants in MODY 1-14 genes among all subjects. The full explanation of detailed variants per patient is displayed in the Supplement Table 1.

**Table 3.**
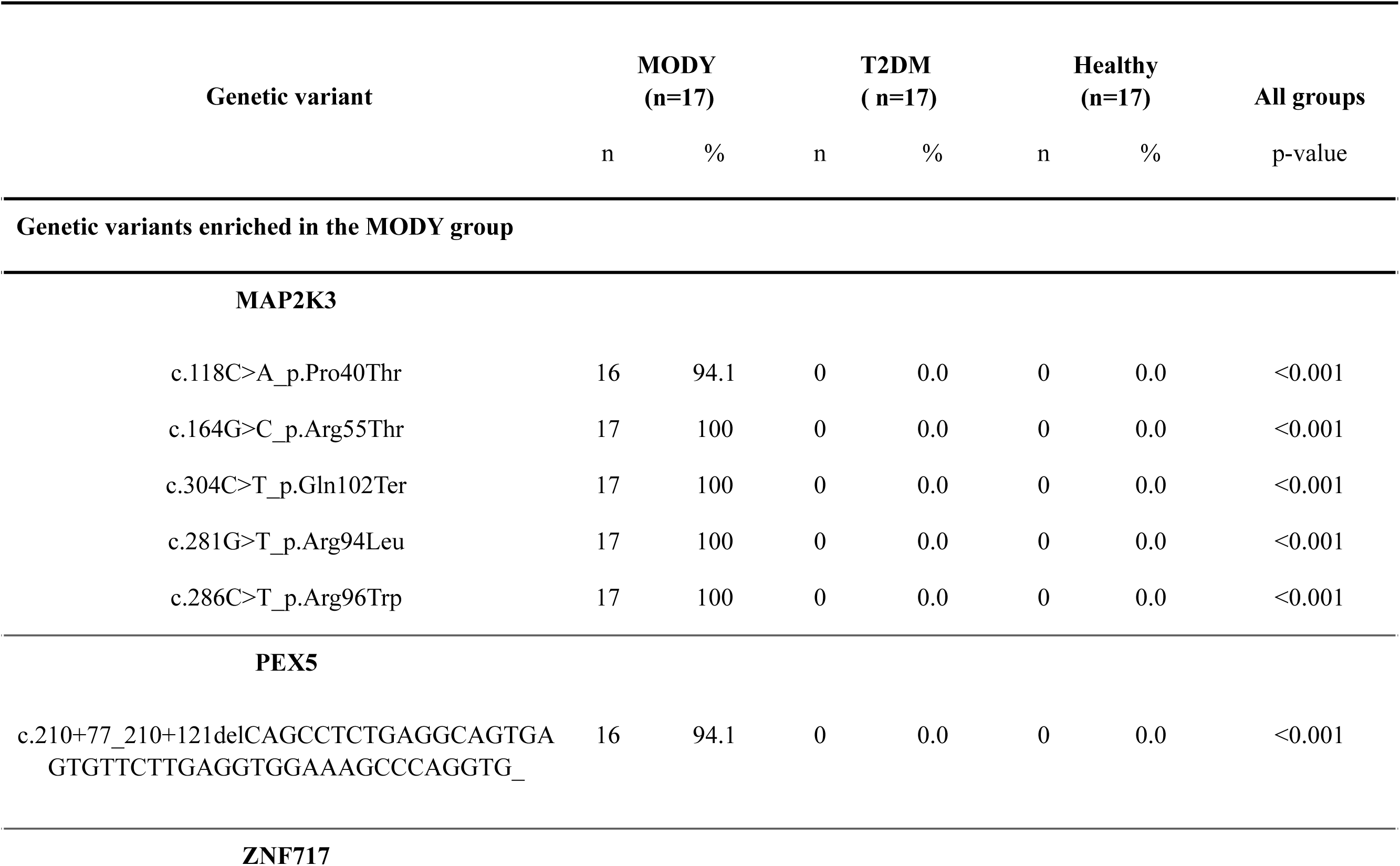
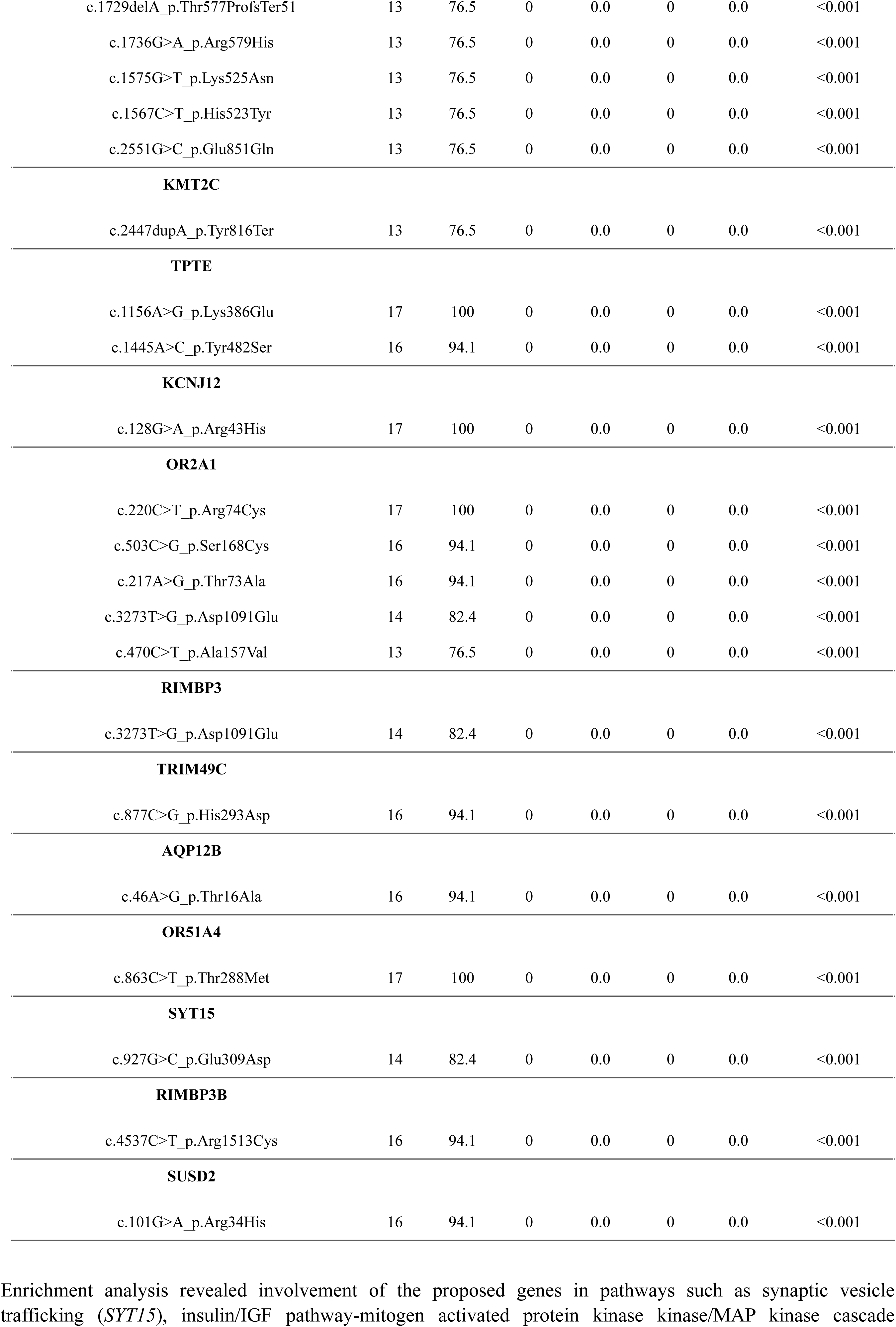
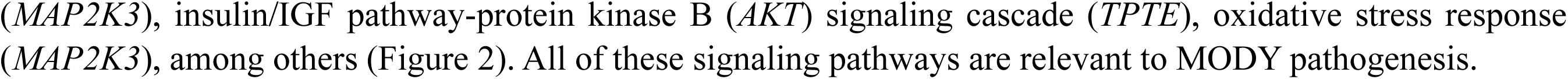
Variant frequency of novel candidate genes with enrichment in the MODY group.

Several *MAP2K3* genetic variants were present in 100% of MODY cases but in none of the patients with T2DM or HC. These variants include c.281G>T (p.Arg94Leu), c.286C>T (p.Arg96Trp), c.304C>T (p.Gln102Termination) and c.164G>C (p.Arg55Thr); and the variant c.164G>C (p.Arg55Thr), found in 94% of MODY cases while in 0% of the rest.

Regarding *TPTE* gene, the variants c.1156A>G (p.Lys386Glu) and c.1445A>C (p.Tyr482Ser) were found in 100% and 94% of MODY cases, respectively while they were absent in T2DM and HC. The c.128G>A (p.Arg43His) variant of the *KCNJ12* gene, was found in 100% of MODY cases but in none of the patients with T2DM or HC. Additional variants found in 100% of MODY cases while absent in T2DM and HC included *OR51A4* c.863C>T_p.Thr288Met, *OR2A1* c.220C>T_p.Arg74Cys, and *SYT15* c.927G>C (p.Glu309Asp)

Additionally, in 94% of cases with MODY but none of the patients with T2DM or HC, we identified a splicing site variant in *PEX5* gene, two additional variants in *OR2A1*, and a single variant in *AQP12B* (c.46A>G_p.Thr16Ala), as well as single variants in *TRIM49C*, *RIMBP3B*, and *SUSD2*.

## Discussion

We have demonstrated that genetic variants in the 14 classical MODY genes cannot differentiate Mexican patients with MODY from patients with T2DM and healthy subjects as they do in Caucasian populations. Only 5% of these genetic variants displayed differences across groups, including the c.C1226T:p.T409I variant in *CEL* which was detected in 58% of patients with MODY but in none of the HC or subjects with T2DM. Patients with MODY presented a high degree of allelic heterogeneity when analyzed individually as they all displayed different variants among these 14 canonical genes, supporting the notion that in some populations this condition behaves in a polygenic manner. Several of our patients with T2DM, on the other hand, carried genetic variants in known MODY genes. In a recent GWAS meta-analysis comprising over 51,000 patients with T2DM, genetic variants in MODY genes *HNF4A, GCK* and *HNF1A*, were found to be associated with a 4-to 8-fold increased diabetes risk.^13^ Taken together, this information represents a paradigm shift in the pathophysiology of both, MODY and T2DM in non-Caucasian populations.

The absence of variants of clinical significance in MODY canonical genes in most of our patients, and the fact that these genes were mainly described in Caucasian populations, prompted us to investigate the presence of other genetic variants that could explain the MODY genetic landscape in our Mexican population and serve as potential diagnostic biomarkers. This knowledge gap is supported by several authors who have pointed to the existence of other genes involved in early onset diabetes in non-Caucasian populations, and need of WES studies to elucidate new genes involved in glycemic regulation. ^7–8, 14–18^ Thus, concomitant variants could contribute to MODY phenotypical presentation trough the interplay of polygenic backgrounds over isolated variants. Other authors have pointed at MODY polygenic traits given the existence of variants with low-penetrance and limited-pathogenicity.^13,14^ Additionally, studies have suggested to consider non-genetic factors as epigenetic modifications, environmental conditions, and ethnicity in MODY, especially in the context of patients who fulfill diagnostic criteria but lack a genetic diagnosis.^14^ Examples of low-penetrance variants include *RFX6* protein truncating variants in Finnish MODY patients without confirmatory diagnosis.^15^

Additional efforts conducted worldwide to better characterize MODY have found other genes and variants linked to this disease. Patel et al. reported the involvement of *RFX6* p.His293Leu variant in Finnish MODY cases.^15^ Genes such as *WFS1*, *NKX6-1,* and *AKT2* have been associated with MODY in South Indian patients^16^. Jakiel et al. proposed candidate genes including *MTOR, TBC1D4, CACNA1E,* and *MNX1* in Polish MODY patients without a genetic diagnosis, and Simaite et al. described the causal role of *PCBD1* mutations in early-onset non-autoimmune diabetes.^17–18^ We intentionally searched for genetic variants in the aforementioned genes and did not find any statistically significant differences among groups.

In the context of a highly heterogeneous genetic disease where current evidence and large-scale genomic studies have been predominantly generated in Caucasian populations, this is the first study to conduct WES in a MODY Latino population, analyzing beyond its canonical genes. Latin America represents a key context to study diabetes genetics due to the disproportionately heavy disease burden present in Latino populations, yet Latin-American patients remain underrepresented in DNA-based studies.^10,11^ We propose 14 candidate genes which are significantly enriched in Mexican patients with MODY and not in subjects with T2DM or HC of the same ethnogenetic background. These variants could serve as diagnostic biomarkers to better identify MODY patients, differentiating them from those with T2DM.

Enrichment analysis revealed the proposed genes to be involved in a myriad of metabolic and signaling pathways such as synaptic vesicle trafficking (*SYT15* gene), hypoxia response via Hypoxia Inducible Factor (HIF) activation, insulin/IGF pathway-mitogen activated protein kinase kinase/MAP kinase cascade (*MAP2K3* gene), insulin/IGF pathway-protein kinase B signaling cascade (*TPTE* gene), and oxidative stress response. Several of the proposed genes have critical implications on glucose metabolism and beta-cell function. In-depth understanding and analysis of the association of each candidate gene represents a key aspect in which our findings may contribute to elucidate MODY pathophysiology and glycemic regulation mechanisms (Figure 3).

**Figure 3.**
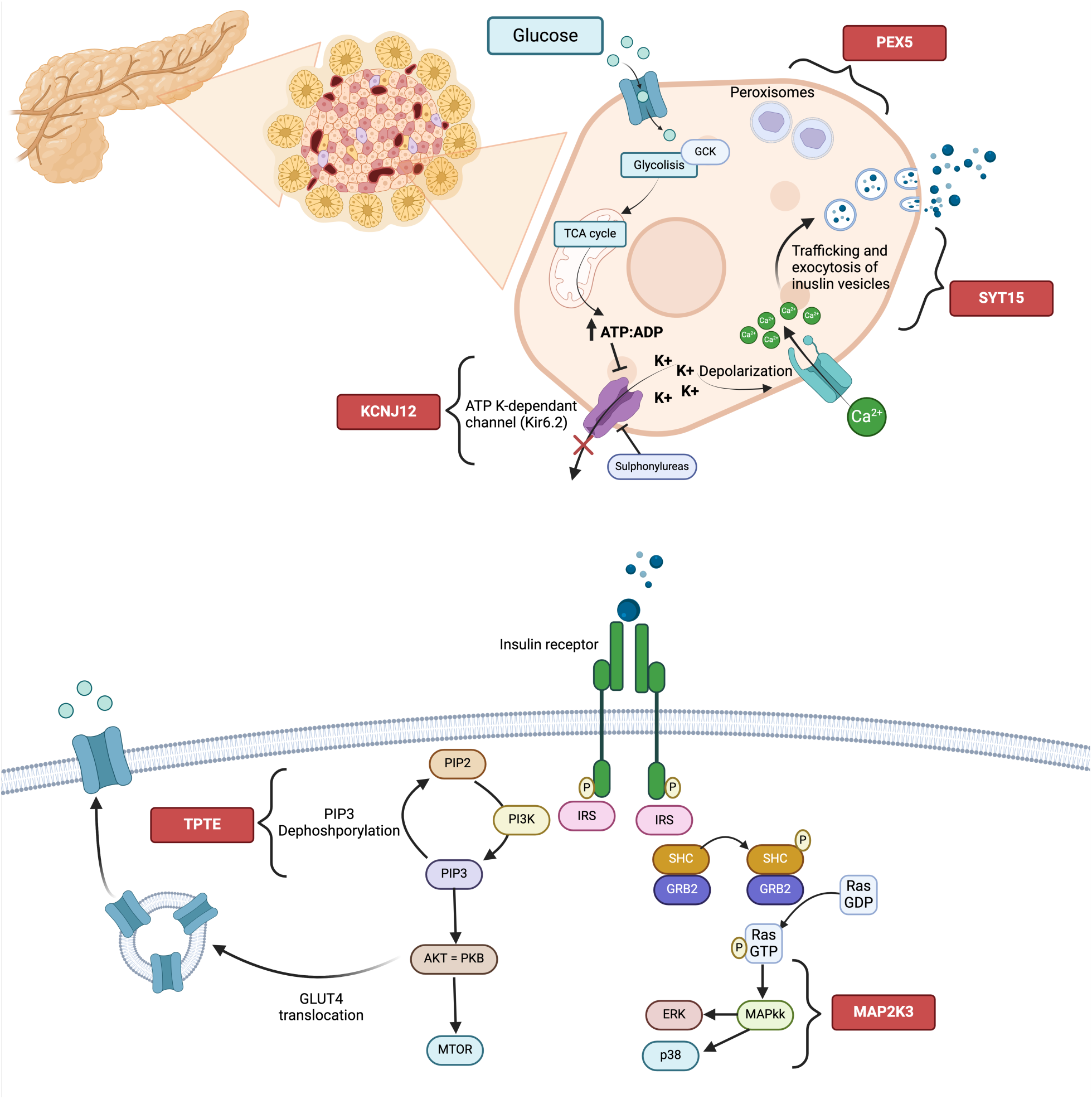
Potential sites of involvement of candidate genes in insulin release and glucose metabolism pathways.

Physiologically, after preproinsulin is synthesized in the endoplasmic reticulum of pancreatic beta-cells, proinsulin is directed to the Golgi apparatus and packaged into secretory granules.^19^ Beta-cell glucose metabolism increases the ATP:ADP ratio, which deactivates the ATP-sensitive K-channels.^20^ Increased intracellular potassium induces membrane depolarization and an increase of calcium influx through the voltage gated channel, promoting vesicle transport and fusion with the plasma membrane to release insulin via exocytosis. Sulfonylureas inhibit the ATP-sensitive potassium channels, which are conformed by the Kir6.2 and sulfonylurea receptor proteins.^20^ In peripheral tissues such as muscle, insulin binds to the insulin receptor (IR), inducing recruitment of insulin receptor substrate (IRS) to phosphorylate the tyrosine kinase domain of the IR and activate two main pathways: the PI3K-Akt and the MAPK.^20^ Firstly, the active IRS activates phosphoinositide 3-kinase (PI3K), which phosphorylates phosphatidylinositol 4,5-bisphosphate (PIP_2_) into phosphatidylinositol 3,4,5-triphosphate (PIP_3_) to activate downstream effectors as AKT/PKB, and mTOR to ultimately promote GLUT4 translocation into the plasmatic membrane.^20^ Second, IRS phosphorylates and activates GRB2/SHC/SOS to induce the phosphorylation of Ras-GDP to Ras-GTP. Then, downstream effectors as MAP kinase kinase (MAPkk) activate ERK1/2 to promote GLUT-4 transcription.^20^

As previously mentioned, synaptic vesicle trafficking is crucial for insulin release. The *SYT15* gene encodes for a membrane trafficking protein belonging to the Synaptotagmin family.^21^ Synaptotagmins are calcium sensors that regulate granule fusion to the plasma membrane.^22^ Synaptotagmin-7 is essential for glucose-stimulated insulin secretion. Gustavsson et al. demonstrated that synaptotagmin-7 null mutant mice present altered glucose-induced insulin secretion, suggesting a calcium-sensing defect^21^. Other Synaptotagmin family members as Synaptotagmin-4 modulate beta-cell maturation, calcium sensitivity, and vesicle release.^23^

The transmembrane phosphatase with tensin homolog (*TPTE*) gene encodes for a PTEN-related tyrosine phosphatase with key roles in endocrine and testicular signal transduction pathways^21^. In insulin signaling pathway, PTEN (Phosphatase and tensin homolog) exerts negative regulation, as dephosphorylates PIP_3_ into PIP_2_ in the PI3K pathway, reducing insulin response.^20^ Thus, loss-of-function mutations increase insulin signaling, especially in clinical contexts such as Cowden syndrome. Nevertheless, other studies have reported PTEN polymorphisms that increase insulin resistance and metabolic syndrome features.^20^

The *KCNJ12* gene, similarly to *KCNJ11* (MODY 13 causal gene) encodes for a potassium inwardly rectifying channel (Kir 2.2).^21^ As previously mentioned, inhibition of potassium influx is crucial for beta-cell depolarization and insulin secretion. In oncogenesis, *KCNJ12* acts as a target for miR-132-3p to modulate the AKT signaling pathway, involved in glucose metabolism by increasing GLUT-1 and GLUT-4 translocation.^24^

The *MAP2K3* gene encodes for a protein that belongs to the MAPkk family, which mediates signaling through phosphorylation of MAP14/p38-MAPK.^21^ In glycemic regulation, MAP2K3 activity is enhanced by insulin, which is necessary for the proper expression of glucose transporters.^25^ Li et al examined the effect of AMPK (AMP-activated protein kinase) in p38-MAPK activation, finding p38-MAPK involvement in glucose transport, while its inhibition reduced glucose uptake and GLUT-4 translocation.^26^ Other authors described altered p38-MAPK activity in T2DM patients’ skeletal muscle, adipose tissue and kidney.^27^

*PEX5* gene plays a crucial role in peroxisomal protein import, as its product binds to the C-terminal PTS1-type tripeptide peroxisomal targeting signal.^21^ PEX5 delivers folded proteins from the cytosol into peroxisomes, translocating them across the membrane and then returning them to the cytosol.^28^ Rip-Pex5 −/− mice present impaired insulin secretion, and glucose intolerance.^28^ Peroxisomes are ubiquitous organelles deeply involved in lipid metabolism, and beta-cell homeostasis. Peroxisome proliferator-activated receptor (PPAR) alpha agonists also confer protection to beta-cells against fatty acids.^29^

To the best of our knowledge, this study represents the first effort to characterize the genomic landscape of MODY patients through WES in Latin America. We propose a novel set of candidate genes involved in MODY diagnosis. The strengths of our study include conducting WES as part of the methodology unlike other studies of MODY using panels with a limited number of genes and the comparison of our findings in MODY patients with HC and subjects with T2DM of the same ethnogenetic background. Limitations include need for external validation of candidate genes, lack of non-coding region analysis and genetic evaluation of MODY patient relatives.

Future perspectives include performing external validation of the candidate genes, functional analysis to strengthen associations, genotype-phenotype association analysis, and development of AI-assisted models to combine diagnostic biomarkers and create polygenic risk scores. The latter could incorporate new tools into clinical practice for a correct diagnosis of MODY in Latino populations and improve patients’ outcomes with individually tailored therapies for MODY patients.

## Data Availability

All data produced in the present study are available upon reasonable request to the authors

## Acknowledgments

None.

## Author Contributions

A.F.H., D.M.R, K.T.P, A.M.N., and M.M. were involved in the conception, and design of the study. F.M.M, S.V.P., E.S.L.A., S.H.A., R.C.S., J.H.P., A.M.N., and I.R.R. contributed to the data collection. S.A.E., A.F.H., D.M.R, A.M.N, K.T.P, and M.M conducted the analysis and interpretation of the results. A.M.N., A.F.H., M.M., E.S.L.A, S.M.M., K.L.W., and R.M.I wrote the first draft of the manuscript, and all authors edited, reviewed, and approved the final version of the manuscript.

## Data and Resource Availability Statement

The datasets generated during and/or analyzed in the current study are available from the corresponding author upon reasonable request.

