## Supplement Table 1 and 2 for "Maturity Onset Diabetes of the Young (MODY) is not always monogenic: candidate genes involved in a Latino Population"

**Table of Contents**

**Supplement Table 1.** Variants in MODY 1-14 genes per patient

**Supplement Table 2.** Frequency of genetic variants among other genes associated with MODY (RFX6 WFS1, NKX6-1, AKT2, NKX2-2, PCBD1, MTOR, TBC1D4, CACNA1E, MNX1).

**Supplement Table 1. Variants in MODY 1-14 genes per patient.**

| Patient #<br>Treatment<br><br>(Suspected MODY) | Pathogenic variants present | Variant type | MODY freq | (%) | T2D freq | (%) | Healthy freq | (%) | P-val All | Exeter score |
|---|---|---|---|---|---|---|---|---|---|---|
| Pt. 1<br>Insulin<br><br>(MODY 3 HNF1A) | HNF1A c.C51G:p.L17L | S | 12 | 70.6 | 14 | 82.4 | 12 | 70.6 | 0.662 | 45.5 |
| | HNF1A c.A79C:p.I27L | NS | 11 | 64.7 | 11 | 64.7 | 10 | 58.8 | 0.92 | |
| | HNF1A c.G864C:p.G288G | S | 9 | 52.9 | 14 | 82.4 | 15 | 88.2 | **0.041** | |
| | HNF1A c.A1741G:p.S581G | NS | 17 | 100 | 17 | 100 | 17 | 100 | NA | |
| | HNF1B c.C606G:p.N202K | NS | 2 | 11.8 | 0 | 0.0 | 0 | 0.0 | 0.125 | |
| | NEUROD1 c.A133G:p.T45A | NS | 16 | 94.1 | 17 | 100 | 16 | 94.1 | 0.346 | |
| | KLF11 c.A1185T:p.V395V | S | 17 | 100 | 17 | 100 | 17 | 100 | NA | |
| | CEL c.C1710T:p.P570P | S | 3 | 17.6 | 9 | 52.9 | 9 | 52.9 | 0.054 | |
| | CEL c.C1164T:p.T388T | S | 3 | 17.6 | 0 | 0.0 | 1 | 5.9 | 0.15 | |
| | CEL c.C2064G:p.G688G | S | 3 | 17.6 | 0 | 0.0 | 0 | 0.0 | **0.041** | |
| | CEL c.C1226T:p.T409I | NS | 10 | 58.8 | 0 | 0.0 | 0 | 0.0 | **<0.001** | |
| | CEL c.T2059G:p.S687A | NS | 2 | 11.8 | 0 | 0.0 | 0 | 0.0 | 0.125 | |
| | CEL c.G2065C:p.A689P | NS | 2 | 11.8 | 0 | 0.0 | 0 | 0.0 | 0.125 | |
| | PAX4 c.A1046G:p.X349W | SL | 16 | 94.1 | 17 | 100 | 17 | 100 | 0.361 | |
| | PAX4 c.A986C:p.H329P | NS | 16 | 94.1 | 17 | 100 | 17 | 100 | 0.361 | |
| | BLK c.T843C:p.F281F | S | 17 | 100 | 16 | 94.1 | 17 | 100 | 0.361 | |
| | BLK c.T330C:p.S110S | S | 8 | 47.1 | 7 | 41.2 | 8 | 47.1 | 0.924 | |
| | ABCC8 c.T207C:p.P69P | S | 16 | 94.1 | 16 | 94.1 | 14 | 82.4 | 0.412 | |
| | ABCC8 c.G4102T:p.A1368S | NS | 14 | 82.4 | 13 | 76.5 | 12 | 70.6 | 0.721 | |
| | ABCC8 c.C1683T:p.H561H | S | 12 | 70.6 | 4 | 23.5 | 11 | 64.7 | **0.011** | |
| | ABCC8 c.G1944A:p.K648K | S | 4 | 23.5 | 1 | 5.9 | 2 | 11.8 | 0.314 | |
| | KCNJ11 c.C309T:p.A103A | S | 5 | 29.4 | 3 | 17.6 | 2 | 11.8 | 0.419 | |
| | KCNJ11 c.G748A:p.V250I | NS | 14 | 82.4 | 13 | 76.5 | 12 | 70.6 | 0.721 | |

| | | | | | | | | | | |
|---|---|---|---|---|---|---|---|---|---|---|
| | KCNJ11 c.A67G:p.K23E | NS | 13 | 76.5 | 13 | 76.5 | 12 | 70.6 | 0.902 | |
| | APPL1 c.A2099G:p.E700G | NS | 4 | 23.5 | 2 | 11.8 | 4 | 23.5 | 0.608 | |
| Pt 2.<br><br>Insulin<br>Metformin<br>Glimepiride<br><br>(MODY 3<br>HNF1A /<br>MODY 5<br>HNF1B) | HNF4A c.C341T:p.T114I | NS | 4 | 23.5 | 1 | 5.9 | 0 | 0.0 | 0.056 | 75.5 |
| | HNF1A c.C51G:p.L17L | S | 12 | 70.6 | 14 | 82.4 | 12 | 70.6 | 0.662 | |
| | HNF1A c.A79C:p.I27L | NS | 11 | 64.7 | 11 | 64.7 | 10 | 58.8 | 0.92 | |
| | HNF1A c.C1375T:p.L459L | S | 9 | 52.9 | 14 | 82.4 | 15 | 88.2 | **0.041** | |
| | HNF1A c.G1460A:p.S487N | NS | 11 | 64.7 | 10 | 58.8 | 8 | 47.1 | 0.571 | |
| | HNF1A c.A1741G:p.S581G | NS | 17 | 100 | 17 | 100 | 17 | 100 | NA | |
| | NEUROD1 c.A133G:p.T45A | NS | 16 | 94.1 | 17 | 100 | 16 | 94.1 | 0.346 | |
| | KLF11 c.A1185T:p.V395V | S | 17 | 100 | 17 | 100 | 17 | 100 | NA | |
| | PAX4 c.A1046G:p.X349W | SL | 16 | 94.1 | 17 | 100 | 17 | 100 | 0.361 | |
| | PAX4 c.A986C:p.H329P | NS | 16 | 94.1 | 17 | 100 | 17 | 100 | 0.361 | |
| | PAX4 c.A543G:p.Q181Q | S | 1 | 5.9 | 2 | 11.8 | 2 | 11.8 | 0.801 | |
| | BLK c.T843C:p.F281F | S | 17 | 100 | 16 | 94.1 | 17 | 100 | 0.361 | |
| | ABCC8 c.G4102T:p.A1368S | NS | 14 | 82.4 | 13 | 76.5 | 12 | 70.6 | 0.721 | |
| | ABCC8 c.G3816A:p.R1272R | S | 10 | 58.8 | 7 | 41.2 | 7 | 41.2 | 0.492 | |
| | ABCC8 c.C1683T:p.H561H | S | 12 | 70.6 | 4 | 23.5 | 11 | 64.7 | **0.011** | |
| | KCNJ11 c.G748A:p.V250I | NS | 14 | 82.4 | 13 | 76.5 | 12 | 70.6 | 0.721 | |
| | KCNJ11 c.A67G:p.K23E | NS | 13 | 76.5 | 13 | 76.5 | 12 | 70.6 | 0.902 | |
| | APPL1 c.A69G:p.L23L | S | 2 | 11.8 | 1 | 5.9 | 0 | 0.0 | 0.346 | |
| Pt. 3<br><br>Insulin<br>Metformin<br>Dapagliflozin<br>Liraglutide<br><br>(MODY 3<br>HNF1A /<br>MODY 5<br>HNF1B) | GCK c.C642T:p.Y214Y | S | 2 | 11.8 | 0 | 0.0 | 0 | 0.0 | 0.125 | 75.5 |
| | HNF1A c.G864C:p.G288G | S | 9 | 52.9 | 14 | 82.4 | 15 | 88.2 | **0.041** | |
| | HNF1A c.A1741G:p.S581G | NS | 17 | 100 | 17 | 100 | 17 | 100 | NA | |
| | NEUROD1 c.A133G:p.T45A | NS | 16 | 94.1 | 17 | 100 | 16 | 94.1 | 0.346 | |
| | KLF11 c.A1185T:p.V395V | S | 17 | 100 | 17 | 100 | 17 | 100 | NA | |
| | CEL c.C1226T:p.T409I | NS | 10 | 58.8 | 0 | 0.0 | 0 | 0.0 | **<0.001** | |
| | BLK c.T843C:p.F281F | S | 17 | 100 | 16 | 94.1 | 17 | 100 | 0.361 | |
| | ABCC8 c.T207C:p.P69P | S | 16 | 94.1 | 16 | 94.1 | 14 | 82.4 | 0.412 | |
| | ABCC8 c.G4102T:p.A1368S | NS | 14 | 82.4 | 13 | 76.5 | 12 | 70.6 | 0.721 | |
| | ABCC8 c.G3816A:p.R1272R | S | 10 | 58.8 | 7 | 41.2 | 7 | 41.2 | 0.492 | |
| | ABCC8 c.C1683T:p.H561H | S | 12 | 70.6 | 4 | 23.5 | 11 | 64.7 | **0.011** | |
| | ABCC8 c.G1720A:p.V574M | NS | 1 | 5.9 | 0 | 0.0 | 0 | 0.0 | 0.361 | |
| | KCNJ11 c.G748A:p.V250I | NS | 14 | 82.4 | 13 | 76.5 | 12 | 70.6 | 0.721 | |
| | APPL1 c.T256C:p.L86L | S | 2 | 11.8 | 0 | 0.0 | 0 | 0.0 | 0.125 | |

| | | | | | | | | | | |
|---|---|---|---|---|---|---|---|---|---|---|
| Pt. 4 Metformin (MODY 3 HNF1A) | GCK c.T668C:p.M223T | NS | 1 | 5.9 | 0 | 0.0 | 0 | 0.0 | 0.361 | 75.5 |
| | HNF1A c.C51G:p.L17L | S | 12 | 70.6 | 14 | 82.4 | 12 | 70.6 | 0.662 | |
| | HNF1A c.A79C:p.I27L | NS | 11 | 64.7 | 11 | 64.7 | 10 | 58.8 | 0.92 | |
| | HNF1A c.C1375T:p.L459L | S | 9 | 52.9 | 14 | 82.4 | 15 | 88.2 | **0.041** | |
| | HNF1A c.G1460A:p.S487N | NS | 11 | 64.7 | 10 | 58.8 | 8 | 47.1 | 0.571 | |
| | HNF1A c.A1741G:p.S581G | NS | 17 | 100 | 17 | 100 | 17 | 100 | NA | |
| | KLF11 c.A1185T:p.V395V | S | 17 | 100 | 17 | 100 | 17 | 100 | NA | |
| | CEL c.C1226T:p.T409I | NS | 10 | 58.8 | 0 | 0.0 | 0 | 0.0 | **<0.001** | |
| | PAX4 c.A1046G:p.X349W | SL | 16 | 94.1 | 17 | 100 | 17 | 100 | 0.361 | |
| | PAX4 c.A986C:p.H329P | NS | 16 | 94.1 | 17 | 100 | 17 | 100 | 0.361 | |
| | BLK c.T843C:p.F281F | S | 17 | 100 | 16 | 94.1 | 17 | 100 | 0.361 | |
| | BLK c.A974C:p.K325T | NS | 1 | 5.9 | 0 | 0.0 | 1 | 5.9 | 0.594 | |
| | ABCC8 c.T207C:p.P69P | S | 16 | 94.1 | 16 | 94.1 | 14 | 82.4 | 0.412 | |
| | ABCC8 c.G4102T:p.A1368S | NS | 14 | 82.4 | 13 | 76.5 | 12 | 70.6 | 0.721 | |
| | ABCC8 c.G3816A:p.R1272R | S | 10 | 58.8 | 7 | 41.2 | 7 | 41.2 | 0.492 | |
| | ABCC8 c.C3609T:p.A1203A | S | 7 | 41.2 | 3 | 17.6 | 5 | 29.4 | 0.322 | |
| | ABCC8 c.C1683T:p.H561H | S | 12 | 70.6 | 4 | 23.5 | 11 | 64.7 | **0.011** | |
| | ABCC8 c.G1944A:p.K648K | S | 4 | 23.5 | 1 | 5.9 | 2 | 11.8 | 0.314 | |
| | KCNJ11 c.G748A:p.V250I | NS | 14 | 82.4 | 13 | 76.5 | 12 | 70.6 | 0.721 | |
| | KCNJ11 c.A67G:p.K23E | NS | 13 | 76.5 | 13 | 76.5 | 12 | 70.6 | 0.902 | |
| | KCNJ11 c.C309T:p.A103A | S | 5 | 29.4 | 3 | 17.6 | 2 | 11.8 | 0.419 | |
| | APPL1 c.A69G:p.L23L | S | 2 | 11.8 | 1 | 5.9 | 0 | 0.0 | 0.346 | |
| Pt. 5 Metformin Linagliptin (MODY 9 PAX4) | HNF1A c.G864C:p.G288G | S | 9 | 52.9 | 14 | 82.4 | 15 | 88.2 | **0.041** | 45.5 |
| | HNF1A c.A1741G:p.S581G | NS | 17 | 100 | 17 | 100 | 17 | 100 | NA | |
| | PDX1 c.A338G:p.N113S | NS | 1 | 5.9 | 0 | 0.0 | 0 | 0.0 | 0.361 | |
| | NEUROD1 c.A133G:p.T45A | NS | 16 | 94.1 | 17 | 100 | 16 | 94.1 | 0.346 | |
| | KLF11 c.A1185T:p.V395V | S | 17 | 100 | 17 | 100 | 17 | 100 | NA | |
| | PAX4 c.A1046G:p.X349W | SL | 16 | 94.1 | 17 | 100 | 17 | 100 | 0.361 | |
| | PAX4 c.A986C:p.H329P | NS | 16 | 94.1 | 17 | 100 | 17 | 100 | 0.361 | |
| | BLK c.T843C:p.F281F | S | 17 | 100 | 16 | 94.1 | 17 | 100 | 0.361 | |
| | BLK c.T330C:p.S110S | S | 8 | 47.1 | 7 | 41.2 | 8 | 47.1 | 0.924 | |
| | ABCC8 c.T207C:p.P69P | S | 16 | 94.1 | 16 | 94.1 | 14 | 82.4 | 0.412 | |

| | | | | | | | | | | |
|---|---|---|---|---|---|---|---|---|---|---|
| | ABCC8 c.C1683T:p.H561H | S | 12 | 70.6 | 4 | 23.5 | 11 | 64.7 | **0.011** | |
| Pt. 6 Metformin Glimepiride (MODY 5 HNF1B) | HNF1A c.C51G:p.L17L | S | 12 | 70.6 | 14 | 82.4 | 12 | 70.6 | 0.662 | 62.4 |
| | HNF1A c.A79C:p.I27L | NS | 11 | 64.7 | 11 | 64.7 | 10 | 58.8 | 0.92 | |
| | HNF1A c.G864C:p.G288G | S | 9 | 52.9 | 14 | 82.4 | 15 | 88.2 | **0.041** | |
| | HNF1A c.C1375T:p.L459L | S | 9 | 52.9 | 14 | 82.4 | 15 | 88.2 | **0.041** | |
| | HNF1A c.G1460A:p.S487N | NS | 11 | 64.7 | 10 | 58.8 | 8 | 47.1 | 0.571 | |
| | HNF1A c.A1741G:p.S581G | NS | 17 | 100 | 17 | 100 | 17 | 100 | NA | |
| | NEUROD1 c.A133G:p.T45A | NS | 16 | 94.1 | 17 | 100 | 16 | 94.1 | 0.346 | |
| | KLF11 c.A1185T:p.V395V | S | 17 | 100 | 17 | 100 | 17 | 100 | NA | |
| | CEL c.C1710T:p.P570P | S | 3 | 17.6 | 9 | 52.9 | 9 | 52.9 | 0.054 | |
| | CEL c.C1164T:p.T388T | S | 3 | 17.6 | 0 | 0.0 | 1 | 5.9 | 0.15 | |
| | CEL c.C2064G:p.G688G | S | 3 | 17.6 | 0 | 0.0 | 0 | 0.0 | **0.041** | |
| | CEL c.C603T:p.F201F | S | 1 | 5.9 | 0 | 0.0 | 0 | 0.0 | 0.361 | |
| | CEL c.C1226T:p.T409I | NS | 10 | 58.8 | 0 | 0.0 | 0 | 0.0 | **<0.001** | |
| | CEL c.T2059G:p.S687A | NS | 2 | 11.8 | 0 | 0.0 | 0 | 0.0 | 0.125 | |
| | CEL c.G2065C:p.A689P | NS | 2 | 11.8 | 0 | 0.0 | 0 | 0.0 | 0.125 | |
| | CEL c.G1801C:p.A601P | NS | 1 | 5.9 | 0 | 0.0 | 0 | 0.0 | 0.361 | |
| | PAX4 c.A1046G:p.X349W | SL | 16 | 94.1 | 17 | 100 | 17 | 100 | 0.361 | |
| | PAX4 c.A986C:p.H329P | NS | 16 | 94.1 | 17 | 100 | 17 | 100 | 0.361 | |
| | BLK c.T843C:p.F281F | S | 17 | 100 | 16 | 94.1 | 17 | 100 | 0.361 | |
| | BLK c.T330C:p.S110S | S | 8 | 47.1 | 7 | 41.2 | 8 | 47.1 | 0.924 | |
| | ABCC8 c.T207C:p.P69P | S | 16 | 94.1 | 16 | 94.1 | 14 | 82.4 | 0.412 | |
| | ABCC8 c.C1683T:p.H561H | S | 12 | 70.6 | 4 | 23.5 | 11 | 64.7 | **0.011** | |
| Pt. 7 Semaglutide Pioglitazone Glimepiride (MODY 3 HNF1A) | HNF4A c.C341T:p.T114I | NS | 4 | 23.5 | 1 | 5.9 | 0 | 0.0 | 0.056 | 75.5 |
| | HNF1A c.C51G:p.L17L | S | 12 | 70.6 | 14 | 82.4 | 12 | 70.6 | 0.662 | |
| | HNF1A c.A1741G:p.S581G | NS | 17 | 100 | 17 | 100 | 17 | 100 | NA | |
| | NEUROD1 c.A133G:p.T45A | NS | 16 | 94.1 | 17 | 100 | 16 | 94.1 | 0.346 | |
| | KLF11 c.A1185T:p.V395V | S | 17 | 100 | 17 | 100 | 17 | 100 | NA | |
| | PAX4 c.A1046G:p.X349W | SL | 16 | 94.1 | 17 | 100 | 17 | 100 | 0.361 | |
| | PAX4 c.A986C:p.H329P | NS | 16 | 94.1 | 17 | 100 | 17 | 100 | 0.361 | |
| | PAX4 c.G680A:p.R227Q | NS | 1 | 5.9 | 0 | 0.0 | 0 | 0.0 | 0.361 | |

| | | | | | | | | | | |
|---|---|---|---|---|---|---|---|---|---|---|
| | BLK c.T843C:p.F281F | S | 17 | 100 | 16 | 94.1 | 17 | 100 | 0.361 | |
| | ABCC8 c.T207C:p.P69P | S | 16 | 94.1 | 16 | 94.1 | 14 | 82.4 | 0.412 | |
| | ABCC8 c.G4102T:p.A1368S | NS | 14 | 82.4 | 13 | 76.5 | 12 | 70.6 | 0.721 | |
| | ABCC8 c.G3816A:p.R1272R | S | 10 | 58.8 | 7 | 41.2 | 7 | 41.2 | 0.492 | |
| | ABCC8 c.C3609T:p.A1203A | S | 7 | 41.2 | 3 | 17.6 | 5 | 29.4 | 0.322 | |
| | ABCC8 c.C1683T:p.H561H | S | 12 | 70.6 | 4 | 23.5 | 11 | 64.7 | **0.011** | |
| | ABCC8 c.C2997T:p.C999C | S | 1 | 5.9 | 0 | 0.0 | 1 | 5.9 | 0.594 | |
| | KCNJ11 c.G748A:p.V250I | NS | 14 | 82.4 | 13 | 76.5 | 12 | 70.6 | 0.721 | |
| | KCNJ11 c.A67G:p.K23E | NS | 13 | 76.5 | 13 | 76.5 | 12 | 70.6 | 0.902 | |
| | KCNJ11 c.C540G:p.L180L | S | 1 | 5.9 | 0 | 0.0 | 0 | 0.0 | 0.361 | |
| Pt. 8 Diet and exercise (MODY 9 PAX4) | GCK c.C642T:p.Y214Y | S | 2 | 11.8 | 0 | 0.0 | 0 | 0.0 | 0.125 | 75.5 |
| | HNF1A c.G864C:p.G288G | S | 9 | 52.9 | 14 | 82.4 | 15 | 88.2 | **0.041** | |
| | HNF1A c.A1741G:p.S581G | NS | 17 | 100 | 17 | 100 | 17 | 100 | NA | |
| | NEUROD1 c.A133G:p.T45A | NS | 16 | 94.1 | 17 | 100 | 16 | 94.1 | 0.346 | |
| | KLF11 c.A185G:p.Q62R | NS | 3 | 17.6 | 1 | 5.9 | 2 | 11.8 | 0.567 | |
| | KLF11 c.A1185T:p.V395V | S | 17 | 100 | 17 | 100 | 17 | 100 | NA | |
| | PAX4 c.A1046G:p.X349W | SL | 16 | 94.1 | 17 | 100 | 17 | 100 | 0.361 | |
| | PAX4 c.A986C:p.H329P | NS | 16 | 94.1 | 17 | 100 | 17 | 100 | 0.361 | |
| | BLK c.T843C:p.F281F | S | 17 | 100 | 16 | 94.1 | 17 | 100 | 0.361 | |
| | BLK c.T330C:p.S110S | S | 8 | 47.1 | 7 | 41.2 | 8 | 47.1 | 0.924 | |
| | ABCC8 c.T207C:p.P69P | S | 16 | 94.1 | 16 | 94.1 | 14 | 82.4 | 0.412 | |
| | ABCC8 c.G4102T:p.A1368S | NS | 14 | 82.4 | 13 | 76.5 | 12 | 70.6 | 0.721 | |
| | ABCC8 c.G3816A:p.R1272R | S | 10 | 58.8 | 7 | 41.2 | 7 | 41.2 | 0.492 | |
| | ABCC8 c.C3609T:p.A1203A | S | 7 | 41.2 | 3 | 17.6 | 5 | 29.4 | 0.322 | |
| | KCNJ11 c.G748A:p.V250I | NS | 14 | 82.4 | 13 | 76.5 | 12 | 70.6 | 0.721 | |
| | KCNJ11 c.A67G:p.K23E | NS | 13 | 76.5 | 13 | 76.5 | 12 | 70.6 | 0.902 | |
| | APPL1 c.A2099G:p.E700G | NS | 4 | 23.5 | 2 | 11.8 | 4 | 23.5 | 0.608 | |
| Pt. 9 Liraglutide Metformin Glimepiride (MODY 3 HNF1A) | HNF1A c.C51G:p.L17L | S | 12 | 70.6 | 14 | 82.4 | 12 | 70.6 | 0.662 | 35.8 |
| | HNF1A c.A79C:p.I27L | NS | 11 | 64.7 | 11 | 64.7 | 10 | 58.8 | 0.92 | |
| | HNF1A c.G864C:p.G288G | S | 9 | 52.9 | 14 | 82.4 | 15 | 88.2 | **0.041** | |
| | HNF1A c.C1375T:p.L459L | S | 9 | 52.9 | 14 | 82.4 | 15 | 88.2 | **0.041** | |
| | HNF1A c.G1460A:p.S487N | NS | 11 | 64.7 | 10 | 58.8 | 8 | 47.1 | 0.571 | |
| | HNF1A c.A1741G:p.S581G | NS | 17 | 100 | 17 | 100 | 17 | 100 | NA | |
| | NEUROD1 c.A133G:p.T45A | NS | 16 | 94.1 | 17 | 100 | 16 | 94.1 | 0.346 | |

| | | | | | | | | | | |
|---|---|---|---|---|---|---|---|---|---|---|
| | KLF11 c.A1185T:p.V395V | S | 17 | 100 | 17 | 100 | 17 | 100 | NA | |
| | CEL c.C1226T:p.T409I | NS | 10 | 58.8 | 0 | 0.0 | 0 | 0.0 | **<0.001** | |
| | PAX4 c.A1046G:p.X349W | SL | 16 | 94.1 | 17 | 100 | 17 | 100 | 0.361 | |
| | PAX4 c.A986C:p.H329P | NS | 16 | 94.1 | 17 | 100 | 17 | 100 | 0.361 | |
| | BLK c.T843C:p.F281F | S | 17 | 100 | 16 | 94.1 | 17 | 100 | 0.361 | |
| | ABCC8 c.T207C:p.P69P | S | 16 | 94.1 | 16 | 94.1 | 14 | 82.4 | 0.412 | |
| | ABCC8 c.G4102T:p.A1368S | NS | 14 | 82.4 | 13 | 76.5 | 12 | 70.6 | 0.721 | |
| | KCNJ11 c.G748A:p.V250I | NS | 14 | 82.4 | 13 | 76.5 | 12 | 70.6 | 0.721 | |
| | KCNJ11 c.A67G:p.K23E | NS | 13 | 76.5 | 13 | 76.5 | 12 | 70.6 | 0.902 | |
| | KCNJ11 c.C309T:p.A103A | S | 5 | 29.4 | 3 | 17.6 | 2 | 11.8 | 0.419 | |
| Pt. 10 Metformin (MODY 3 HNF1A) | HNF1A c.C51G:p.L17L | S | 12 | 70.6 | 14 | 82.4 | 12 | 70.6 | 0.662 | 75.5 |
| | HNF1A c.A79C:p.I27L | NS | 11 | 64.7 | 11 | 64.7 | 10 | 58.8 | 0.92 | |
| | HNF1A c.G864C:p.G288G | S | 9 | 52.9 | 14 | 82.4 | 15 | 88.2 | **0.041** | |
| | HNF1A c.C1375T:p.L459L | S | 9 | 52.9 | 14 | 82.4 | 15 | 88.2 | **0.041** | |
| | HNF1A c.G1460A:p.S487N | NS | 11 | 64.7 | 10 | 58.8 | 8 | 47.1 | 0.571 | |
| | HNF1A c.A1741G:p.S581G | NS | 17 | 100 | 17 | 100 | 17 | 100 | NA | |
| | NEUROD1 c.A133G:p.T45A | NS | 16 | 94.1 | 17 | 100 | 16 | 94.1 | 0.346 | |
| | KLF11 c.A1185T:p.V395V | S | 17 | 100 | 17 | 100 | 17 | 100 | NA | |
| | KLF11 c.828_829insTCTGTC: p.V280_P281insSV | NF ins | 1 | 5.9 | 0 | 0.0 | 0 | 0.0 | 0.361 | |
| | PAX4 c.A1046G:p.X349W | SL | 16 | 94.1 | 17 | 100 | 17 | 100 | 0.361 | |
| | PAX4 c.A986C:p.H329P | NS | 16 | 94.1 | 17 | 100 | 17 | 100 | 0.361 | |
| | BLK c.T843C:p.F281F | S | 17 | 100 | 16 | 94.1 | 17 | 100 | 0.361 | |
| | ABCC8 c.T207C:p.P69P | S | 16 | 94.1 | 16 | 94.1 | 14 | 82.4 | 0.412 | |
| | ABCC8 c.G4102T:p.A1368S | NS | 14 | 82.4 | 13 | 76.5 | 12 | 70.6 | 0.721 | |
| | ABCC8 c.G3816A:p.R1272R | S | 10 | 58.8 | 7 | 41.2 | 7 | 41.2 | 0.492 | |
| | ABCC8 c.C3609T:p.A1203A | S | 7 | 41.2 | 3 | 17.6 | 5 | 29.4 | 0.322 | |
| | KCNJ11 c.G748A:p.V250I | NS | 14 | 82.4 | 13 | 76.5 | 12 | 70.6 | 0.721 | |
| | KCNJ11 c.A67G:p.K23E | NS | 13 | 76.5 | 13 | 76.5 | 12 | 70.6 | 0.902 | |
| | APPL1 c.A2099G:p.E700G | NS | 4 | 23.5 | 2 | 11.8 | 4 | 23.5 | 0.608 | |

| | | | | | | | | | | |
|---|---|---|---|---|---|---|---|---|---|---|
| Pt. 11 Insulin (MODY 3 HNF1A) | HNF1A c.C51G:p.L17L | S | 12 | 70.6 | 14 | 82.4 | 12 | 70.6 | 0.662 | 45.5 |
| | HNF1A c.A79C:p.I27L | NS | 11 | 64.7 | 11 | 64.7 | 10 | 58.8 | 0.92 | |
| | HNF1A c.C1375T:p.L459L | S | 9 | 52.9 | 14 | 82.4 | 15 | 88.2 | **0.041** | |
| | HNF1A c.G1460A:p.S487N | NS | 11 | 64.7 | 10 | 58.8 | 8 | 47.1 | 0.571 | |
| | HNF1A c.A1741G:p.S581G | NS | 17 | 100 | 17 | 100 | 17 | 100 | NA | |
| | NEUROD1 c.A133G:p.T45A | NS | 16 | 94.1 | 17 | 100 | 16 | 94.1 | 0.346 | |
| | KLF11 c.A1185T:p.V395V | S | 17 | 100 | 17 | 100 | 17 | 100 | NA | |
| | CEL c.C1226T:p.T409I | NS | 10 | 58.8 | 0 | 0.0 | 0 | 0.0 | **<0.001** | |
| | CEL c.G41C:p.C14S | NS | 1 | 5.9 | 0 | 0.0 | 0 | 0.0 | 0.361 | |
| | CEL c.T1454C:p.I485T | NS | 1 | 5.9 | 0 | 0.0 | 0 | 0.0 | 0.361 | |
| | PAX4 c.A1046G:p.X349W | SL | 16 | 94.1 | 17 | 100 | 17 | 100 | 0.361 | |
| | PAX4 c.A986C:p.H329P | NS | 16 | 94.1 | 17 | 100 | 17 | 100 | 0.361 | |
| | BLK c.T843C:p.F281F | S | 17 | 100 | 16 | 94.1 | 17 | 100 | 0.361 | |
| | BLK c.T330C:p.S110S | S | 8 | 47.1 | 7 | 41.2 | 8 | 47.1 | 0.924 | |
| | BLK c.C570T:p.S190S | S | 1 | 5.9 | 0 | 0.0 | 0 | 0.0 | 0.361 | |
| | ABCC8 c.T207C:p.P69P | S | 16 | 94.1 | 16 | 94.1 | 14 | 82.4 | 0.412 | |
| | ABCC8 c.G4102T:p.A1368S | NS | 14 | 82.4 | 13 | 76.5 | 12 | 70.6 | 0.721 | |
| | ABCC8 c.G3816A:p.R1272R | S | 10 | 58.8 | 7 | 41.2 | 7 | 41.2 | 0.492 | |
| | ABCC8 c.C3609T:p.A1203A | S | 7 | 41.2 | 3 | 17.6 | 5 | 29.4 | 0.322 | |
| | ABCC8 c.C1683T:p.H561H | S | 12 | 70.6 | 4 | 23.5 | 11 | 64.7 | **0.011** | |
| | ABCC8 c.G1944A:p.K648K | S | 4 | 23.5 | 1 | 5.9 | 2 | 11.8 | 0.314 | |
| | KCNJ11 c.G748A:p.V250I | NS | 14 | 82.4 | 13 | 76.5 | 12 | 70.6 | 0.721 | |
| | KCNJ11 c.A67G:p.K23E | NS | 13 | 76.5 | 13 | 76.5 | 12 | 70.6 | 0.902 | |
| Pt. 12 Insulin (MODY 2 GCK) | HNF1A c.C51G:p.L17L | S | 12 | 70.6 | 14 | 82.4 | 12 | 70.6 | 0.662 | 75.5 |
| | HNF1A c.A79C:p.I27L | NS | 11 | 64.7 | 11 | 64.7 | 10 | 58.8 | 0.92 | |
| | HNF1A c.C1375T:p.L459L | S | 9 | 52.9 | 14 | 82.4 | 15 | 88.2 | **0.041** | |
| | HNF1A c.G1460A:p.S487N | NS | 11 | 64.7 | 10 | 58.8 | 8 | 47.1 | 0.571 | |
| | HNF1A c.A1741G:p.S581G | NS | 17 | 100 | 17 | 100 | 17 | 100 | NA | |
| | NEUROD1 c.A133G:p.T45A | NS | 16 | 94.1 | 17 | 100 | 16 | 94.1 | 0.346 | |
| | KLF11 c.A1185T:p.V395V | S | 17 | 100 | 17 | 100 | 17 | 100 | NA | |
| | CEL c.G2021A:p.G674D | NS | 1 | 5.9 | 0 | 0.0 | 0 | 0.0 | 0.361 | |

| | | | | | | | | | | |
|---|---|---|---|---|---|---|---|---|---|---|
| | PAX4 c.A1046G:p.X349W | SL | 16 | 94.1 | 17 | 100 | 17 | 100 | 0.361 | |
| | PAX4 c.A986C:p.H329P | NS | 16 | 94.1 | 17 | 100 | 17 | 100 | 0.361 | |
| | BLK c.T843C:p.F281F | S | 17 | 100 | 16 | 94.1 | 17 | 100 | 0.361 | |
| | ABCC8 c.T207C:p.P69P | S | 16 | 94.1 | 16 | 94.1 | 14 | 82.4 | 0.412 | |
| | ABCC8 c.G4102T:p.A1368S | NS | 14 | 82.4 | 13 | 76.5 | 12 | 70.6 | 0.721 | |
| | ABCC8 c.C2482T:p.L828L | S | 1 | 5.9 | 2 | 11.8 | 3 | 17.6 | 0.567 | |
| | ABCC8 c.C1683T:p.H561H | S | 12 | 70.6 | 4 | 23.5 | 11 | 64.7 | **0.011** | |
| | ABCC8 c.C2274T:p.T758T | S | 1 | 5.9 | 0 | 0.0 | 0 | 0.0 | 0.361 | |
| | KCNJ11 c.G748A:p.V250I | NS | 14 | 82.4 | 13 | 76.5 | 12 | 70.6 | 0.721 | |
| | KCNJ11 c.A67G:p.K23E | NS | 13 | 76.5 | 13 | 76.5 | 12 | 70.6 | 0.902 | |
| | KCNJ11 c.C309T:p.A103A | S | 5 | 29.4 | 3 | 17.6 | 2 | 11.8 | 0.419 | |
| Pt. 13 | HNF4A c.C341T:p.T114I | NS | 4 | 23.5 | 1 | 5.9 | 0 | 0.0 | 0.056 | 75.5 |
| Insulin Linagliptin | HNF1A c.C51G:p.L17L | S | 12 | 70.6 | 14 | 82.4 | 12 | 70.6 | 0.662 | |
| (MODY 3 HNF1A / MODY 5 HNF1B) | HNF1A c.A79C:p.I27L | NS | 11 | 64.7 | 11 | 64.7 | 10 | 58.8 | 0.92 | |
| | HNF1A c.G864C:p.G288G | S | 9 | 52.9 | 14 | 82.4 | 15 | 88.2 | **0.041** | |
| | HNF1A c.C1375T:p.L459L | S | 9 | 52.9 | 14 | 82.4 | 15 | 88.2 | **0.041** | |
| | HNF1A c.G1460A:p.S487N | NS | 11 | 64.7 | 10 | 58.8 | 8 | 47.1 | 0.571 | |
| | HNF1A c.A1741G:p.S581G | NS | 17 | 100 | 17 | 100 | 17 | 100 | NA | |
| | NEUROD1 c.A133G:p.T45A | NS | 16 | 94.1 | 17 | 100 | 16 | 94.1 | 0.346 | |
| | KLF11 c.A185G:p.Q62R | NS | 3 | 17.6 | 1 | 5.9 | 2 | 11.8 | 0.567 | |
| | KLF11 c.A1185T:p.V395V | S | 17 | 100 | 17 | 100 | 17 | 100 | NA | |
| | CEL c.C1226T:p.T409I | NS | 10 | 58.8 | 0 | 0.0 | 0 | 0.0 | **<0.001** | |
| | PAX4 c.A1046G:p.X349W | SL | 16 | 94.1 | 17 | 100 | 17 | 100 | 0.361 | |
| | PAX4 c.A986C:p.H329P | NS | 16 | 94.1 | 17 | 100 | 17 | 100 | 0.361 | |
| | BLK c.T843C:p.F281F | S | 17 | 100 | 16 | 94.1 | 17 | 100 | 0.361 | |
| | BLK c.T330C:p.S110S | S | 8 | 47.1 | 7 | 41.2 | 8 | 47.1 | 0.924 | |
| | BLK c.C711T:p.P237P | S | 1 | 5.9 | 0 | 0.0 | 0 | 0.0 | 0.361 | |
| | ABCC8 c.T207C:p.P69P | S | 16 | 94.1 | 16 | 94.1 | 14 | 82.4 | 0.412 | |
| | ABCC8 c.G4102T:p.A1368S | NS | 14 | 82.4 | 13 | 76.5 | 12 | 70.6 | 0.721 | |

| | | | | | | | | | | |
|---|---|---|---|---|---|---|---|---|---|---|
| | KCNJ11 c.G748A:p.V250I | NS | 14 | 82.4 | 13 | 76.5 | 12 | 70.6 | 0.721 | |
| | KCNJ11 c.A67G:p.K23E | NS | 13 | 76.5 | 13 | 76.5 | 12 | 70.6 | 0.902 | |
| Pt. 14 | HNF1A c.C51G:p.L17L | S | 12 | 70.6 | 14 | 82.4 | 12 | 70.6 | 0.662 | 58.0 |
| Dapagliflozin Metformin Glimepiride | HNF1A c.A79C:p.I27L | NS | 11 | 64.7 | 11 | 64.7 | 10 | 58.8 | 0.92 | |
| | HNF1A c.C1375T:p.L459L | S | 9 | 52.9 | 14 | 82.4 | 15 | 88.2 | **0.041** | |
| (MODY 3 HNF1A / MODY 5 HNF1B) | HNF1A c.G1460A:p.S487N | NS | 11 | 64.7 | 10 | 58.8 | 8 | 47.1 | 0.571 | |
| | HNF1A c.A1741G:p.S581G | NS | 17 | 100 | 17 | 100 | 17 | 100 | NA | |
| | NEUROD1 c.A133G:p.T45A | NS | 16 | 94.1 | 17 | 100 | 16 | 94.1 | 0.346 | |
| | KLF11 c.A185G:p.Q62R | NS | 3 | 17.6 | 1 | 5.9 | 2 | 11.8 | 0.567 | |
| | KLF11 c.A1185T:p.V395V | S | 17 | 100 | 17 | 100 | 17 | 100 | NA | |
| | CEL c.C1226T:p.T409I | NS | 10 | 58.8 | 0 | 0.0 | 0 | 0.0 | **<0.001** | |
| | CEL c.2032dupC:p.V681Rfs*6 | F ins | 1 | 5.9 | 0 | 0.0 | 0 | 0.0 | 0.361 | |
| | PAX4 c.A1046G:p.X349W | SL | 16 | 94.1 | 17 | 100 | 17 | 100 | 0.361 | |
| | PAX4 c.A986C:p.H329P | NS | 16 | 94.1 | 17 | 100 | 17 | 100 | 0.361 | |
| | BLK c.T843C:p.F281F | S | 17 | 100 | 16 | 94.1 | 17 | 100 | 0.361 | |
| | ABCC8 c.T207C:p.P69P | S | 16 | 94.1 | 16 | 94.1 | 14 | 82.4 | 0.412 | |
| | ABCC8 c.G4102T:p.A1368S | NS | 14 | 82.4 | 13 | 76.5 | 12 | 70.6 | 0.721 | |
| | ABCC8 c.G3816A:p.R1272R | S | 10 | 58.8 | 7 | 41.2 | 7 | 41.2 | 0.492 | |
| | ABCC8 c.C3609T:p.A1203A | S | 7 | 41.2 | 3 | 17.6 | 5 | 29.4 | 0.322 | |
| | KCNJ11 c.G748A:p.V250I | NS | 14 | 82.4 | 13 | 76.5 | 12 | 70.6 | 0.721 | |
| | KCNJ11 c.A67G:p.K23E | NS | 13 | 76.5 | 13 | 76.5 | 12 | 70.6 | 0.902 | |
| Pt. 15 | HNF1A c.G864C:p.G288G | S | 9 | 52.9 | 14 | 82.4 | 15 | 88.2 | **0.041** | 75.5 |
| Metformin Glimepiride Linagliptin Dapagliflozin | HNF1A c.A1741G:p.S581G | NS | 17 | 100 | 17 | 100 | 17 | 100 | NA | |
| | NEUROD1 c.A133G:p.T45A | NS | 16 | 94.1 | 17 | 100 | 16 | 94.1 | 0.346 | |
| (MODY 3 HNF1A) | KLF11 c.A1185T:p.V395V | S | 17 | 100 | 17 | 100 | 17 | 100 | NA | |
| | CEL c.C1226T:p.T409I | NS | 10 | 58.8 | 0 | 0.0 | 0 | 0.0 | **<0.001** | |
| | PAX4 c.A1046G:p.X349W | SL | 16 | 94.1 | 17 | 100 | 17 | 100 | 0.361 | |
| | PAX4 c.A986C:p.H329P | NS | 16 | 94.1 | 17 | 100 | 17 | 100 | 0.361 | |
| | BLK c.T843C:p.F281F | S | 17 | 100 | 16 | 94.1 | 17 | 100 | 0.361 | |
| | ABCC8 c.T207C:p.P69P | S | 16 | 94.1 | 16 | 94.1 | 14 | 82.4 | 0.412 | |
| | ABCC8 c.G4102T:p.A1368S | NS | 14 | 82.4 | 13 | 76.5 | 12 | 70.6 | 0.721 | |
| | ABCC8 c.G3816A:p.R1272R | S | 10 | 58.8 | 7 | 41.2 | 7 | 41.2 | 0.492 | |

| | | | | | | | | | | |
|---|---|---|---|---|---|---|---|---|---|---|
| | ABCC8 c.C3609T:p.A1203A | S | 7 | 41.2 | 3 | 17.6 | 5 | 29.4 | 0.322 | |
| | ABCC8 c.C1683T:p.H561H | S | 12 | 70.6 | 4 | 23.5 | 11 | 64.7 | **0.011** | |
| | KCNJ11 c.G748A:p.V250I | NS | 14 | 82.4 | 13 | 76.5 | 12 | 70.6 | 0.721 | |
| | KCNJ11 c.A67G:p.K23E | NS | 13 | 76.5 | 13 | 76.5 | 12 | 70.6 | 0.902 | |
| | APPL1 c.A2099G:p.E700G | NS | 4 | 23.5 | 2 | 11.8 | 4 | 23.5 | 0.608 | |
| Pt. 16 Metformin (MODY 3 HNF1A) | HNF1A c.C51G:p.L17L | S | 12 | 70.6 | 14 | 82.4 | 12 | 70.6 | 0.662 | 75.5 |
| | HNF1A c.A79C:p.I27L | NS | 11 | 64.7 | 11 | 64.7 | 10 | 58.8 | 0.92 | |
| | HNF1A c.C1375T:p.L459L | S | 9 | 52.9 | 14 | 82.4 | 15 | 88.2 | **0.041** | |
| | HNF1A c.G1460A:p.S487N | NS | 11 | 64.7 | 10 | 58.8 | 8 | 47.1 | 0.571 | |
| | HNF1A c.A1741G:p.S581G | NS | 17 | 100 | 17 | 100 | 17 | 100 | NA | |
| | HNF1 c.1137delT:p.V380Sfs*4 | F ins | 1 | 5.9 | 0 | 0.0 | 0 | 0.0 | 0.361 | |
| | HNF1B c.C606G:p.N202K | NS | 2 | 11.8 | 0 | 0.0 | 0 | 0.0 | 0.125 | |
| | NEUROD1 c.A133G:p.T45A | NS | 16 | 94.1 | 17 | 100 | 16 | 94.1 | 0.346 | |
| | KLF11 c.A1185T:p.V395V | S | 17 | 100 | 17 | 100 | 17 | 100 | NA | |
| | CEL c.C1710T:p.P570P | S | 3 | 17.6 | 9 | 52.9 | 9 | 52.9 | 0.054 | |
| | CEL c.C1164T:p.T388T | S | 3 | 17.6 | 0 | 0.0 | 1 | 5.9 | 0.15 | |
| | CEL c.C2064G:p.G688G | S | 3 | 17.6 | 0 | 0.0 | 0 | 0.0 | **0.041** | |
| | CEL c.C1226T:p.T409I | NS | 10 | 58.8 | 0 | 0.0 | 0 | 0.0 | **<0.001** | |
| | PAX4 c.A1046G:p.X349W | SL | 16 | 94.1 | 17 | 100 | 17 | 100 | 0.361 | |
| | PAX4 c.A986C:p.H329P | NS | 16 | 94.1 | 17 | 100 | 17 | 100 | 0.361 | |
| | BLK c.T843C:p.F281F | S | 17 | 100 | 16 | 94.1 | 17 | 100 | 0.361 | |
| | BLK c.T330C:p.S110S | S | 8 | 47.1 | 7 | 41.2 | 8 | 47.1 | 0.924 | |
| | ABCC8 c.T207C:p.P69P | S | 16 | 94.1 | 16 | 94.1 | 14 | 82.4 | 0.412 | |
| | ABCC8 c.G4102T:p.A1368S | NS | 14 | 82.4 | 13 | 76.5 | 12 | 70.6 | 0.721 | |
| | ABCC8 c.G3816A:p.R1272R | S | 10 | 58.8 | 7 | 41.2 | 7 | 41.2 | 0.492 | |
| | ABCC8 c.C1683T:p.H561H | S | 12 | 70.6 | 4 | 23.5 | 11 | 64.7 | **0.011** | |
| | ABCC8 c.G1944A:p.K648K | S | 4 | 23.5 | 1 | 5.9 | 2 | 11.8 | 0.314 | |
| | KCNJ11 c.G748A:p.V250I | NS | 14 | 82.4 | 13 | 76.5 | 12 | 70.6 | 0.721 | |
| | KCNJ11 c.A67G:p.K23E | NS | 13 | 76.5 | 13 | 76.5 | 12 | 70.6 | 0.902 | |
| | KCNJ11 c.C309T:p.A103A | S | 5 | 29.4 | 3 | 17.6 | 2 | 11.8 | 0.419 | |
| | APPL1 c.T256C:p.L86L | S | 2 | 11.8 | 0 | 0.0 | 0 | 0.0 | 0.125 | |

| | | | | | | | | | | |
|---|---|---|---|---|---|---|---|---|---|---|
| Pt. 17 Insulin (MODY 2 GCK / MODY 5 HNF1B) | HNF4A c.C341T:p.T114I | NS | 4 | 23.5 | 1 | 5.9 | 0 | 0.0 | 0.056 | 49.4 |
| | HNF1A c.C1375T:p.L459L | S | 9 | 52.9 | 14 | 82.4 | 15 | 88.2 | **0.041** | |
| | HNF1A c.G1460A:p.S487N | NS | 11 | 64.7 | 10 | 58.8 | 8 | 47.1 | 0.571 | |
| | HNF1A c.A1741G:p.S581G | NS | 17 | 100 | 17 | 100 | 17 | 100 | NA | |
| | NEUROD1 c.A133G:p.T45A | NS | 16 | 94.1 | 17 | 100 | 16 | 94.1 | 0.346 | |
| | KLF11 c.A1185T:p.V395V | S | 17 | 100 | 17 | 100 | 17 | 100 | NA | |
| | PAX4 c.A1046G:p.X349W | SL | 16 | 94.1 | 17 | 100 | 17 | 100 | 0.361 | |
| | PAX4 c.A986C:p.H329P | NS | 16 | 94.1 | 17 | 100 | 17 | 100 | 0.361 | |
| | BLK c.T843C:p.F281F | S | 17 | 100 | 16 | 94.1 | 17 | 100 | 0.361 | |
| | BLK c.T330C:p.S110S | S | 8 | 47.1 | 7 | 41.2 | 8 | 47.1 | 0.924 | |
| | ABCC8 c.T207C:p.P69P | S | 16 | 94.1 | 16 | 94.1 | 14 | 82.4 | 0.412 | |
| | ABCC8 c.C1683T:p.H561H | S | 12 | 70.6 | 4 | 23.5 | 11 | 64.7 | **0.011** | |

We provide the calculated p-value through a chi-square test to compare genetic variant frequencies between all groups. MODY freq = Frequency in the MODY group; T2DM freq = Frequency in the Type 2 Diabetes group; HC Freq = Frequency in the Healthy control group. Pt = Patient; S = synonymous; NS = Non synonymous; NF del = Non frameshift deletion; SL = Stop-loss; F ins = Frameshift insertion; NF ins = Non frameshift insertion; F del = Frameshift deletion; SG = Stop gain.

**Supplement Table 2. Frequency of genetic variants among other genes associated with MODY (RFX6, WFS1, NKX6-1, AKT2, NKX2-2, PCBD1, MTOR, TBC1D4, CACNA1E, MNX1).**

| **Genetic variant** | **MODY (n=17)** | | **T2DM ( n=17)** | | **Healthy (n=17)** | | **All groups** |
|---|---|---|---|---|---|---|---|
| | n | % | n | % | n | % | p-value |
| **RFX6** | | | | | | | |
| c.T1383C:p.T461T | 12 | 70.6 | 11 | 64.7 | 8 | 47.1 | 0.343 |
| c.T1542C:p.N514N | 12 | 70.6 | 11 | 64.7 | 8 | 47.1 | 0.343 |
| c.C1782T:p.H594H | 12 | 70.6 | 11 | 64.7 | 8 | 47.1 | 0.343 |
| c.T1914C:p.G638G | 12 | 70.6 | 11 | 64.7 | 8 | 47.1 | 0.343 |
| c.G985A:p.V329I | 3 | 17.6 | 2 | 11.8 | 2 | 11.8 | 0.847 |
| c.C544T:p.L182F | 0 | 0.0 | 1 | 5.9 | 0 | 0.0 | 0.361 |
| **WFS1** | | | | | | | |
| c.C684G:p.R228R | 16 | 94.1 | 16 | 94.1 | 16 | 94.1 | 1.00 |
| c.G997A:p.V333I | 16 | 94.1 | 17 | 100.0 | 16 | 94.1 | 0.594 |

| | | | | | | | |
|---|---|---|---|---|---|---|---|
| c.C1185T:p.V395V | 15 | 88.2 | 16 | 94.1 | 16 | 94.1 | 0.762 |
| c.C1500T:p.N500N | 15 | 88.2 | 16 | 94.1 | 16 | 94.1 | 0.762 |
| c.G1832A:p.R611H | 15 | 88.2 | 16 | 94.1 | 14 | 82.4 | 0.567 |
| c.G2433A:p.K811K | 15 | 88.2 | 16 | 94.1 | 16 | 94.1 | 0.762 |
| c.A2565G:p.S855S | 16 | 94.1 | 16 | 94.1 | 16 | 94.1 | 1.00 |
| c.G1726A:p.G576S | 1 | 5.9 | 1 | 5.9 | 2 | 11.8 | 0.762 |
| c.C1023T:p.F341F | 2 | 11.8 | 1 | 5.9 | 2 | 11.8 | 0.801 |
| c.C1725T:p.A575A | 2 | 11.8 | 0 | 0.0 | 2 | 11.8 | 0.338 |
| c.G2322A:p.K774K | 1 | 5.9 | 0 | 0.0 | 2 | 11.8 | 0.346 |
| c.G1367A:p.R456H | 2 | 11.8 | 0 | 0.0 | 2 | 11.8 | 0.338 |
| c.C2666T:p.A889V | 0 | 0.0 | 0 | 0.0 | 1 | 5.9 | 0.361 |
| c.C342T:p.A114A | 0 | 0.0 | 2 | 11.8 | 1 | 5.9 | 0.346 |
| c.G226A:p.G76S | 0 | 0.0 | 0 | 0.0 | 1 | 5.9 | 0.361 |
| c.A2432G:p.K811R | 0 | 0.0 | 0 | 0.0 | 1 | 5.9 | 0.361 |
| c.G817A:p.E273K | 1 | 5.9 | 1 | 5.9 | 0 | 0.0 | 0.594 |
| c.G2623A:p.V875M | 1 | 5.9 | 0 | 0.0 | 0 | 0.0 | 0.361 |
| c.G2596T:p.D866Y | 1 | 5.9 | 0 | 0.0 | 0 | 0.0 | 0.361 |
| c.G2040A:p.E680E | 1 | 5.9 | 0 | 0.0 | 0 | 0.0 | 0.361 |
| **NKX6-1** | | | | | | | |
| c.414_415insTCCTCCGCCTCTGCC:p.A138_A139insSSASA | 1 | 5.9 | 0 | 0.0 | 1 | 5.9 | 0.594 |
| c.G85A:p.A29T | 0 | 0.0 | 0 | 0.0 | 1 | 5.9 | 0.361 |
| c.G462A:p.A154A | 0 | 0.0 | 1 | 5.9 | 0 | 0.0 | 0.361 |
| **AKT2** | | | | | | | |
| c.G1110T:p.P370P | 0 | 0.0 | 1 | 5.9 | 0 | 0.0 | 0.361 |
| **NKX2-2** | | | | | | | |
| c.A64G:p.N22D | 1 | 5.9 | 0 | 0.0 | 0 | 0.0 | 0.361 |
| c.G365C:p.G122A | 1 | 5.9 | 0 | 0.0 | 1 | 5.9 | 0.594 |
| **PCBD1** | | | | | | | |
| No variants found | 0 | 0.0 | 0 | 0.0 | 0 | 0.0 | NA |
| **MTOR** | | | | | | | |

| | | | | | | | |
|---|---|---|---|---|---|---|---|
| c.G4731A:p.A1577A | 15 | 88.2 | 15 | 88.2 | 17 | 100.0 | 0.338 |
| c.C2997T:p.N999N | 15 | 88.2 | 17 | 100.0 | 16 | 94.1 | 0.346 |
| c.T1437C:p.D479D | 15 | 88.2 | 17 | 100.0 | 16 | 94.1 | 0.346 |
| c.T4260C:p.N1420N | 1 | 5.9 | 1 | 5.9 | 0 | 0.0 | 0.594 |
| c.G6909A:p.L2303L | 2 | 11.8 | 3 | 17.6 | 2 | 11.8 | 0.847 |
| c.C5553T:p.S1851S | 2 | 11.8 | 3 | 17.6 | 2 | 11.8 | 0.847 |
| c.C5469T:p.A1823A | 3 | 17.6 | 1 | 5.9 | 1 | 5.9 | 0.412 |
| c.G3462C:p.R1154R | 3 | 17.6 | 1 | 5.9 | 1 | 5.9 | 0.412 |
| c.C4449T:p.C1483C | 0 | 0.0 | 1 | 5.9 | 0 | 0.0 | 0.361 |
| c.T255C:p.G85G | 0 | 0.0 | 1 | 5.9 | 0 | 0.0 | 0.361 |
| c.C6808T:p.R2270W | 0 | 0.0 | 1 | 5.9 | 0 | 0.0 | 0.361 |
| c.A4356G:p.K1452K | 0 | 0.0 | 1 | 5.9 | 0 | 0.0 | 0.361 |
| c.C4376T:p.A1459V | 0 | 0.0 | 0 | 0.0 | 2 | 11.8 | 0.125 |
| c.A5397G:p.E1799E | 0 | 0.0 | 0 | 0.0 | 1 | 5.9 | 0.361 |
| c.C7279T:p.L2427L | 0 | 0.0 | 0 | 0.0 | 1 | 5.9 | 0.361 |
| c.C4556T:p.A1519V | 0 | 0.0 | 0 | 0.0 | 1 | 5.9 | 0.361 |
| c.G6016T:p.V2006F | 0 | 0.0 | 0 | 0.0 | 1 | 5.9 | 0.361 |
| c.A7256G:p.E2419G | 0 | 0.0 | 0 | 0.0 | 1 | 5.9 | 0.361 |
| c.G1148A:p.S383N | 0 | 0.0 | 0 | 0.0 | 1 | 5.9 | 0.361 |
| **TBC1D4** | | | | | | | |
| c.G2455A:p.V819I | 12 | 70.6 | 16 | 94.1 | 16 | 94.1 | 0.071 |
| c.T1611G:p.S537S | 5 | 29.4 | 9 | 52.9 | 5 | 29.4 | 0.261 |
| c.G723C:p.G241G | 17 | 100.0 | 17 | 100.0 | 17 | 100.0 | 1.00 |
| c.C84G:p.P28P | 10 | 58.8 | 8 | 47.1 | 13 | 76.5 | 0.21 |
| c.T3824C:p.V1275A | 1 | 5.9 | 3 | 17.6 | 1 | 5.9 | 0.412 |
| c.C2901T:p.L967L | 12 | 70.6 | 14 | 82.4 | 11 | 64.7 | 0.502 |
| c.C606T:p.F202F | 3 | 17.6 | 1 | 5.9 | 2 | 11.8 | 0.567 |
| c.C3440T:p.T1147M | 1 | 5.9 | 0 | 0.0 | 2 | 11.8 | 0.346 |
| c.C302T:p.A101V | 1 | 5.9 | 2 | 11.8 | 2 | 11.8 | 0.801 |
| c.G2324A:p.R775H | 0 | 0.0 | 1 | 5.9 | 0 | 0.0 | 0.361 |
| c.G1902A:p.P634P | 0 | 0.0 | 1 | 5.9 | 0 | 0.0 | 0.361 |
| c.G330A:p.T110T | 0 | 0.0 | 1 | 5.9 | 0 | 0.0 | 0.361 |

| | | | | | | | |
|---|---|---|---|---|---|---|---|
| c.C1046G:p.S349W | 0 | 0.0 | 0 | 0.0 | 1 | 5.9 | 0.361 |
| c.A2913T:p.G971G | 0 | 0.0 | 0 | 0.0 | 1 | 5.9 | 0.361 |
| c.C3238T:p.P1080S | 0 | 0.0 | 0 | 0.0 | 1 | 5.9 | 0.361 |
| c.A2254G:p.T752A | 0 | 0.0 | 0 | 0.0 | 1 | 5.9 | 0.361 |
| **CACNA1E** | | | | | | | |
| c.T2577A:p.D859E | 4 | 23.5 | 2 | 11.8 | 2 | 11.8 | 0.553 |
| c.G2992A:p.G998S | 1 | 5.9 | 0 | 0.0 | 1 | 5.9 | 0.594 |
| c.C3447T:p.I1149I | 4 | 23.5 | 2 | 11.8 | 2 | 11.8 | 0.553 |
| c.T4008C:p.H1336H | 8 | 47.1 | 7 | 41.2 | 6 | 35.3 | 0.784 |
| c.G5863A:p.A1955T | 8 | 47.1 | 12 | 70.6 | 11 | 64.7 | 0.343 |
| c.C5073T:p.N1691N | 7 | 41.2 | 7 | 41.2 | 6 | 35.3 | 0.921 |
| c.A750G:p.A250A | 1 | 5.9 | 2 | 11.8 | 0 | 0.0 | 0.346 |
| c.C6567T:p.S2189S | 1 | 5.9 | 3 | 17.6 | 1 | 5.9 | 0.412 |
| c.2137_2142del:p.T713_K714del | 0 | 0.0 | 1 | 5.9 | 0 | 0.0 | 0.361 |
| c.C6536T:p.A2179V | 0 | 0.0 | 1 | 5.9 | 0 | 0.0 | 0.361 |
| c.A1935G:p.A645A | 0 | 0.0 | 1 | 5.9 | 0 | 0.0 | 0.361 |
| c.C4905T:p.D1635D | 0 | 0.0 | 1 | 5.9 | 0 | 0.0 | 0.361 |
| c.A2706G:p.G902G | 0 | 0.0 | 0 | 0.0 | 1 | 5.9 | 0.361 |
| c.A3078G:p.P1026P | 0 | 0.0 | 0 | 0.0 | 1 | 5.9 | 0.361 |
| c.G3362A:p.R1121H | 0 | 0.0 | 0 | 0.0 | 1 | 5.9 | 0.361 |
| c.C2260T:p.H754Y | 0 | 0.0 | 0 | 0.0 | 1 | 5.9 | 0.361 |
| c.G6869A:p.G2290E | 0 | 0.0 | 0 | 0.0 | 1 | 5.9 | 0.361 |
| **MNX1** | | | | | | | |
| c.C444T:p.G148G | 0 | 0.0 | 1 | 5.9 | 0 | 0.0 | 0.361 |
| c.G981A:p.E327E | 0 | 0.0 | 1 | 5.9 | 0 | 0.0 | 0.361 |
| c.C47A:p.A16D | 0 | 0.0 | 2 | 11.8 | 0 | 0.0 | 0.125 |
| c.G357T:p.P119P | 0 | 0.0 | 0 | 0.0 | 1 | 5.9 | 0.361 |
| c.T429C:p.P143P | 0 | 0.0 | 0 | 0.0 | 1 | 5.9 | 0.361 |

We provide the calculated p-value through a chi-square test to compare genetic variant frequencies between all groups.
